# COMPASS: A Clinically-Optimized Multimodal Prediction Architecture with Survival Strategy for AD Prognosis

**DOI:** 10.64898/2026.06.23.26356398

**Authors:** Tianqi Liu, Yinuo Wu, Yihang Bao, Wenhao Li, Chaohan Li, Zhe Liu, Guan Ning Lin

## Abstract

Precise early diagnosis and progression prediction of Alzheimer’s Disease (AD) are critical for optimizing clinical intervention. However, current methodologies often suffer from the passive utilization of clinical priors and rigid modal fusion strategies, failing to capture the heterogeneous variations of imaging biomarkers. Furthermore, predicting the precise time-to-conversion from Mild Cognitive Impairment (MCI) to AD remains a formidable challenge. To address these limitations, we propose COMPASS, a clinical-guided multi-modal framework that unifies diagnosis with a comprehensive survival strategy. Specifically, we instigate a paradigm shift to “clinical-prior-driven” learning by incorporating Clinical-Guided Spatial Attention (CGSA), which actively transforms clinical states into visual signals to modulate neural focus on pathological regions. To bridge the semantic gap between modalities, we introduce Reciprocal Semantic Interaction (RSI) via cross-attention, while a Disease-Stage-Aware Modal Fusion (DSAMF) module dynamically adjusts modal weights based on inferred disease severity to mimic clinical reasoning. Moreover, we specifically design a Dual-Head Joint Survival Risk and Time Prediction Network (DH-Net) to jointly perform quantitative conversion time prediction and patient risk stratification. Extensive experiments demonstrate that COMPASS outperforms state-of-the-art methods, achieving 83.19% accuracy in pMCI vs. sMCI classification, an MAE of 7.96 months for conversion time prediction, and a C-index of 0.819. Furthermore, we conducted in-depth neurobiological interpretability analyses, revealing right hippocampal dominance and synergistic regional impairment patterns, thereby providing new biological insights for early AD diagnosis and subtype identification.

## 1. Introduction

Alzheimer’s Disease (AD) is an irreversible neurodegenerative disorder. Current estimates indicate that over 35 million individuals worldwide are affected, with incidence rates rising rapidly [1, 2]. Given that AD precipitates severe cognitive impairment and a loss of independence, coupled with the current absence of effective curative treatments, early screening is of paramount importance [3]. As a prodromal stage of AD, Mild Cognitive Impairment (MCI) represents a transitional phase between Cognitively Normal (CN) aging and dementia, characterized by cognitive decline while daily living activities remain relatively preserved. Based on clinical outcomes within a 36-month window, MCI is categorized into progressive MCI (pMCI) and stable MCI (sMCI) [4]. Considering the high conversion risk associated with pMCI, early and precise identification of these patients, along with the prediction of their specific time-to-conversion, holds significant clinical value for delaying disease progression or potentially reversing cognitive impairment. As illustrated in Fig. 1, in contrast to the distinct structural atrophy observed in AD, pMCI and sMCI exhibit high visual similarity on conventional MRI scans, rendering manual differentiation difficult. This underscores the urgent necessity for developing high-sensitivity computational computer-aided diagnostic tools.

**Figure 1.**
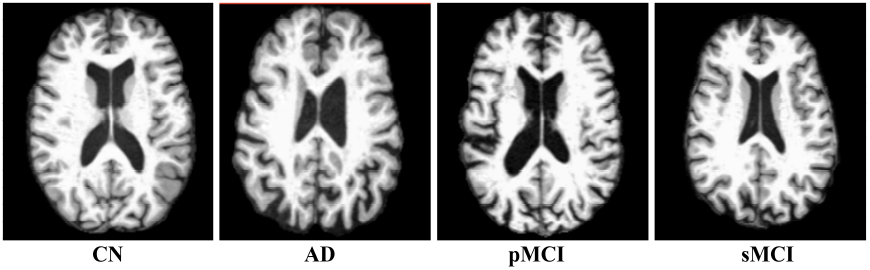
Visualization of brain structural atrophy across distinct stages of AD.

The spatiotemporal heterogeneity of AD pathology renders single-modality approaches insufficient for capturing the comprehensive disease landscape, prompting a paradigm shift towards multi-modal synergistic diagnosis [5]. Among imaging modalities, sMRI and PET are predominant [6]; sMRI quantifies structural atrophy via high-resolution anatomical data [7], while PET sensitively captures early functional anomalies through metabolic levels [8]. Fusing these modalities leverages complementary anatomical and metabolic information, significantly outperforming unimodal approaches [9–12]. Moreover, clinical decision making relies heavily on non-imaging data, such as cognitive scores and genotypes [13–15]. Accordingly, recent models have evolved to integrate multi-source physiological data for holistic patient representation. However, most existing frameworks remain confined to binary classification (e.g., AD vs. CN), largely neglecting the continuous temporal dynamics of disease progression. Although limited studies have addressed survival analysis [16–18], predicting the precise conversion time for pMCI patients remains largely unexplored. To date, a unified framework capable of seamlessly integrating diagnosis, progression prediction, and survival analysis is lacking, representing a critical gap in holistic AD management.

Despite the consensus on multimodal integration, solving the inherent discrepancies in distribution and representation between high-dimensional imaging and low-dimensional non-imaging data remains a formidable challenge. While strategies ranging from input-level early fusion to decision-level late fusion have been explored [13–15, 19, 20], feature-level fusion has emerged as the mainstream approach due to its capacity to extract high-level semantics via non-linear mapping. However, existing frameworks exhibit significant technical limitations regarding the complex pathology of AD. Even in multi-modal approaches, research largely focuses on aligning MRI and PET, while non-imaging data are often integrated via simple feature concatenation. This static strategy of physical stacking fails to bridge the substantial semantic gap between modalities, preventing the establishment of deep intrinsic cross-modal mappings. Furthermore, current models employ fixed fusion weights, ignoring the distinct temporal dynamics of AD where metabolic anomalies typically precede structural atrophy. Consequently, they lack the adaptability to dynamically adjust modal emphasis based on inferred disease severity, failing to simulate clinical reasoning that prioritizes different biomarkers across pathological stages. Critically, the utilization of clinical priors remains passive; in most architectures, clinical data serve merely as auxiliary inputs to terminal layers rather than functioning as contextual cues that actively guide the visual network towards pathological brain regions. This absence of clinical-aware feature extraction not only constrains diagnostic accuracy but also severely compromises model interpretability.

Current methodologies prioritize performance optimization, often overlooking interpretability and physiological significance. Despite achieving state-of-the-art accuracy, the inherent black-box nature of deep learning hinders clinical trust. While some studies have attempted to mitigate this issue using visualization techniques such as Class Activation Mapping (CAM) [21] and Grad-CAM [22], these analyses remain largely confined to qualitative visual demonstrations, lacking deep quantitative exploration of underlying neurobiological mechanisms. Specifically, existing analysis paradigms overlook fine-grained features pivotal to early AD pathology. On one hand, there is a distinct lack of attention to structural lateralization. Neuropsychological evidence suggests that memory impairment in early AD may be associated with specific hemispheric dominance (e.g., the susceptibility of the right hippocampus related to spatial memory [23–25]). However, existing deep learning models rarely perform systematic quantification of hemispheric asymmetry, causing this potential early-stage biomarker to be submerged within global features. On the other hand, network-level insights into synergistic impairment patterns are often absent. AD is fundamentally a network disconnection syndrome [26, 27] rather than isolated regional atrophy; yet, current approaches struggle to disentangle biologically plausible synergistic impairment patterns [28] from learned features. This absence of clinical-pathological validation not only constrains clinician trust in the model’s decision logic but also results in missed opportunities to leverage AI for discovering novel neuroimaging biomarkers.

To address these challenges, we propose COMPASS, the first clinical-guided multi-modal framework integrating diagnosis, conversion prediction, and survival analysis. It incorporates Clinical-Guided Spatial Attention (CGSA) to modulate focus on pathological regions, shifting feature calibration from “data-driven” to “clinical-prior-driven.” To bridge semantic gaps and adapt to disease progression, we introduce Reciprocal Semantic Interaction (RSI) and Disease-Stage-Aware Modal Fusion (DSAMF), establishing deep semantic mappings and mimicking the clinical logic of dynamically adjusting modal emphasis. Operating within a two-stage multi-task framework, COMPASS realizes a holistic workflow for AD management. Furthermore, our analysis reveals the dominant role of the right hippocampus in early AD conversion and identifies synergistic impairment patterns. These findings not only neurobiologically validate the model’s reliability but also provide novel evidence for early AD subtyping and precision medicine.

In summary, the main contributions are as follows:

- We propose COMPASS, the first clinical-guided framework unifying diagnosis, conversion time prediction, and survival analysis, providing comprehensive support for early AD intervention.
- We introduce the CGSA, RSI, and DSAMF modules to align visual representations with clinical priors. These components mimic clinical reasoning by establishing semantic mappings and dynamically weighting biomarkers based on disease progression.
- Extensive experiments demonstrate that COMPASS outperforms State-of-the-Art (SOTA) methods across classification, conversion time prediction, and survival analysis tasks, validating its potential for precision medicine.
- In-depth neurobiological analyses reveal right hippocampal dominance and synergistic impairment patterns in early AD. These insights validate the model’s biological plausibility and offer novel clues for identifying pathological trajectories.

## 2. Related Work

### 2.1. Single-Modality Imaging Methods

Neuroimaging is a pivotal technology for elucidating brain activity and detecting pathological lesions, playing a central role in the clinical diagnosis of AD. Early deep learning research in AD predominantly focused on single-modality analysis using MRI or PET. Based on input dimensionality, these approaches can be categorized into patch-level, slice-level, voxel-level, and subject-level methods. Patch-level methods excel in extracting fine-grained local features, effectively capturing subtle anomalies while suppressing global noise; however, they often overlook holistic spatial correlations. A representative model, sMRI-PatchNet proposed by Zhang et al. [7], utilizes a SHapley Additive exPlanations (SHAP) mechanism to select discriminative patches for classification and atrophy localization. Slice-level methods prioritize computational efficiency and multi-view analysis, aiming to rapidly identify structural variations via 2D slices. To address the loss of 3D spatial information, Chen et al. [29] developed MSA3D, which incorporates an attention mechanism to integrate local and global features across coronal, sagittal, and axial views. Voxel-level methods offer the highest resolution for detecting metabolic or structural anomalies. For instance, Ding et al. [8] designed an end-to-end voxel-level prediction model based on PET images. Despite high accuracy, this approach incurs substantial computational costs. Generally, these three methodologies focus on partial image regions, potentially leading to the loss of critical disease-related information. In contrast, Subject-level methods directly process whole-brain 3D images, offering advantages in global pattern recognition and end-to-end classification. For example, Zhu et al. [30] proposed a deep learning framework that utilizes complete MRI scans as subject-level input, extracting features via 3D Convolutional Neural Network (CNN) and identifying regional anomalies using spatial attention blocks. Similarly, Shou et al. [31] leveraged the informational richness of full MRI scans to disentangle age-related normal atrophy from disease-specific pathological atrophy using deep learning techniques. More recently, Wang et al. [32] utilized whole-brain imaging to construct a global ordinal metric space. By adopting this holistic view, their model employs an exponential moving average strategy to dynamically learn global prototypes for CN and AD subjects, thereby mapping MCI patients onto the trajectory from health to disease.

Overall, subject-based approaches, which extract features from whole-brain scans to exploit global contextual information, have emerged as the dominant input paradigm. However, despite significant advances in feature extraction and classification—particularly with subject-level methods—the performance ceiling of single-modality approaches remains constrained by the physical characteristics and limited information dimensions of a single modality.

### 2.2. Multi-Modality Imaging Methods

In recent years, numerous studies [9–11] have leveraged multi-modal neuroimaging data for AD diagnosis. For instance, Castellano et al. [9] utilized paired MRI and PET data to learn a shared representation by establishing a bi-directional mapping between the original feature space and the latent shared space. Addressing the challenge of missing or mismatched MRI and PET scans, Liu et al. [10] proposed synthesizing the missing modalities to perform AD diagnosis based on completed multi-modal data. Concurrently, various complex architectures have been introduced into multi-modal fusion. Yang et al. [11] employed a Vision Mamba-based self-supervised framework to process large-scale unlabeled multi-modal data, aiming to capture long-range dependencies within images. Similarly, Li et al. [12] proposed the Diamond framework. Built upon the Vision Transformer (ViT) architecture [33], this model separately encodes PET and MRI data and utilizes a dual-attention mechanism to model inter-modal similarities.

In summary, current fusion methods for multi-modal imaging data primarily focus on exploiting novel techniques and frameworks to enhance AD diagnostic performance. However, AD is a complex systemic disease. In real-world practice, clinicians invariably rely on a broader spectrum of information beyond imaging data for diagnosis. Since the pathological evolution of AD is not solely manifested in structural or functional imaging anomalies, incorporating additional non-imaging data often yields significant benefits in capturing comprehensive AD-related pathological information.

### 2.3. Integration of Imaging and Non-Imaging Data

In clinical practice, integrating neuroimaging (e.g., MRI, PET) with non-imaging data (e.g., genetics, cognitive scores) is standard for holistic AD assessment. Consequently, bridging the inherent distributional gap between high-dimensional imaging and low-dimensional non-imaging data has become a primary research focus. Existing strategies generally fall into early, late, and feature-level fusion [13–15, 19, 20]. Early fusion merges raw data at the input layer to capture low-level complementarity—for instance, Mirabnahrazam et al. [19] fused MRI features with genetic SNPs—but often suffers from noise and the curse of dimensionality. Conversely, late fusion aggregates independent predictions at the decision level, such as Mustafa et al.’s [20] ensemble of MRI/CT classifiers. While efficient, this approach inherently neglects deep non-linear interactions between modalities. To address these limitations, feature-level fusion has emerged as the mainstream paradigm, aiming to integrate high-order abstractions within deep neural networks. For example, Zhang et al. [13] employed Transformers to allow non-imaging data to modulate imaging features, enabling deep semantic interaction. Shen et al. [14] proposed TriLightNet, utilizing cascaded attention for efficient, progressive integration, while Yin et al. [15] introduced the UNICORSS framework, which uses clinical metadata to actively calibrate latent feature distributions via contrastive learning.

Despite this progress, significant limitations persist in the utilization of non-imaging data. First, many methods rely excessively on high-cost, hard-to-acquire data (e.g., genomics) to pursue marginal performance gains, neglecting the accessibility of routine clinical variables in real-world scenarios. Second, valuable clinical priors are often utilized passively as terminal auxiliary inputs. This prevents them from serving as contextual cues that actively guide the visual network toward pathological regions. These limitations underscore the urgent need for novel paradigms that efficiently leverage accessible clinical data, transforming clinical priors from passive inputs into active guiding mechanisms.

### 2.4. Survival Analysis in AD Progression

While predominant research has focused on AD diagnosis (i.e., classification tasks), a growing subset of studies is addressing survival analysis within AD progression. This field is currently witnessing a paradigm shift from traditional statistical methodologies to deep learning approaches. Early investigations largely relied on the Cox Proportional Hazards Model [16], utilizing clinical scales and demographic characteristics as covariates. For instance, Barnes et al. [17] developed a point-based risk scoring tool using the Cox model to integrate baseline clinical features (e.g., functional dependency metrics and cognitive scores), enabling the stratification of conversion risk from MCI to probable AD. However, traditional Cox models typically assume a linear relationship between covariates and log-hazards, limiting their capacity to effectively process high-dimensional, dense, and complex neuroimaging data. With the advent of machine learning, researchers have pivoted towards non-parametric and non-linear models to better capture complex interactions and longitudinal dynamics. Musto et al. [18] pioneered the application of Transformer architectures to this domain. Addressing the invasiveness of traditional biomarkers, they utilized blood metabolomics to construct a survival Transformer for precise prognostic stratification. Concurrently, imaging data has been incorporated into survival analysis. Mirabnahrazam et al. [34] employed CNNs to predict progression trajectories by analyzing 63 multi-modal features, including MRI scans, genetics, and clinical data. Similarly, Aghajanian et al. [35] leveraged longitudinal MRI, extracting deep CNN features via 3D Residual Networks (ResNet) and employing Long Short-Term Memory models to assess the risk of MCI-to-AD conversion.

Despite these advancements, existing paradigms suffer from a significant limitation: diagnostic classification and progression prediction are frequently treated as isolated tasks. This disjointed approach neglects the intrinsic correlation between current disease severity (classification features) and future progression velocity (survival features). Consequently, current models fail to capture the fine-grained temporal dynamics of the MCI-to-AD conversion process and struggle to predict precise conversion times. This deficiency severely limits their clinical utility in assisting physicians to formulate precise, time-windowed intervention strategies.

## 3. Method

In this section, we will elaborate on our proposed method, COMPASS, whose overall framework is illustrated in Fig. 2.

**Figure 2.**
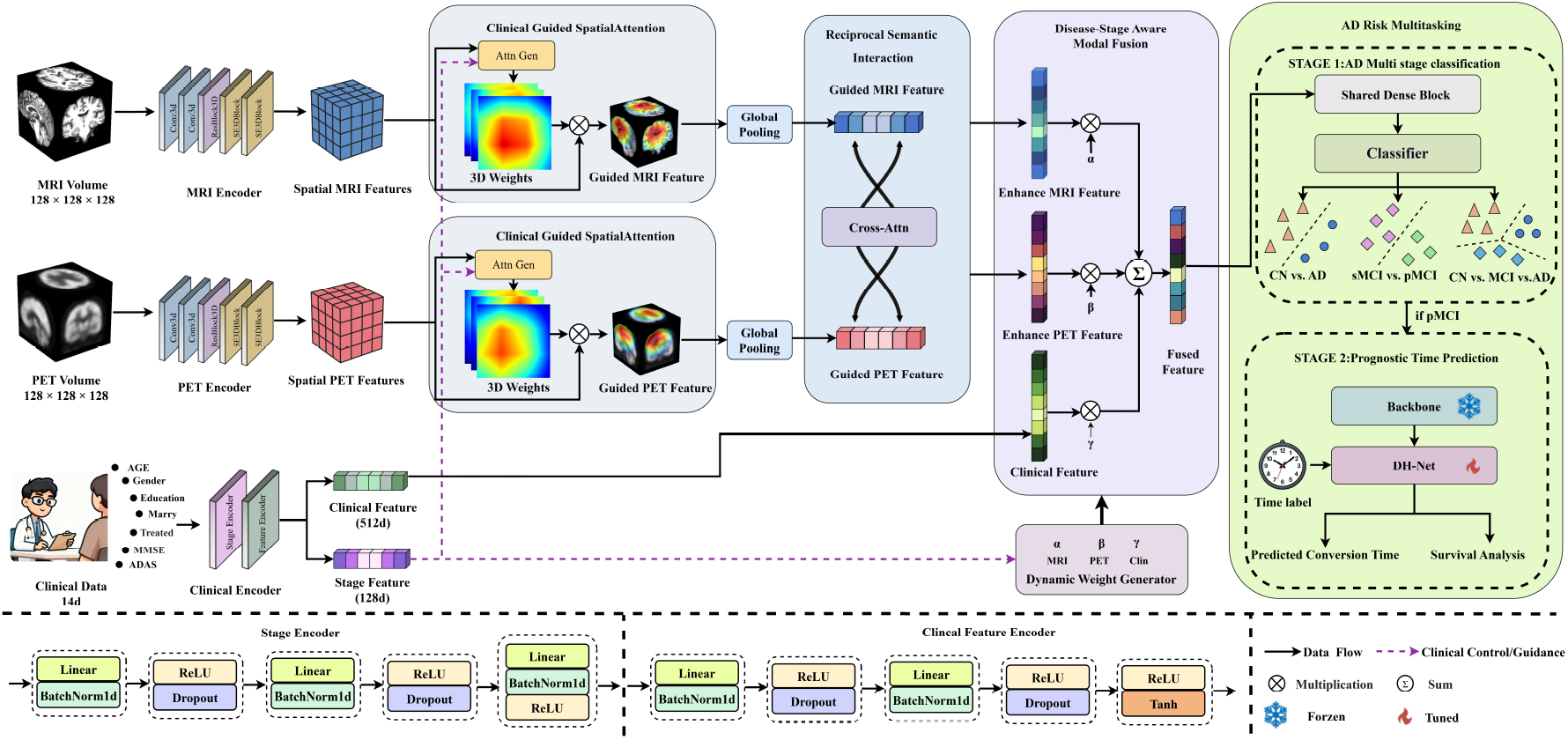
Framework of the COMPASS. Initially, convert MRI/PET volumes and clinical data to spatial/clinical/stage features via corresponding encoders; Then, enhance modal features with clinical-guided spatial attention, fuse bimodal features via reciprocal semantic interaction, and integrate multi-modal features with dynamic weights; Finally, the multi-task module performs AD multi-stage classification, prognostic time prediction, and survival analysis.

### 3.1. Multi-modal Feature Encoder

Constructing an efficient backbone encoder is pivotal for developing a robust AD foundation model. To transform raw tri-modal inputs into high-dimensional semantic representations, we design two specialized encoders dedicated to processing imaging data (MRI, PET) and clinical data, respectively.

#### Imaging Encoder

For 3D medical imaging, diverging from prevalent approaches that employ complex ViT, we opt for a 3D ResNet cite29 enhanced with the Squeeze-and-Excitation (SE) mechanism as the encoder; the architecture is illustrated in Fig. 2. Given an input image **I** ∈ ℝ^1×*D*×*H*×*W*^, the network processes the volume through four cascaded residual stages.

Let **F**^(*l*−1)^ denote the input to the *l*-th residual block. We define the residual mapping function ℱ(·) as a stack of two 3 × 3 × 3 convolutional layers, each followed by Batch Normalization (BN) and ReLU activation. The SE mechanism first acts on this residual mapping to compute channel attention weights **s**:

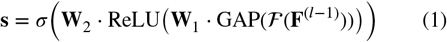

Where GAP(·) denotes global average pooling, *σ*(·) is the Sigmoid function, and **W**_1_, **W**_2_ are the learned weights of the fully connected layers in the SE module. These weights are used to recalibrate the residual features, which are then added to the input shortcut to generate the block output:

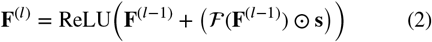

Where ⊙ denotes channel-wise scaling. This formulation explicitly defines how the SE-optimized residual features are integrated with the identity path. We retain the spatial structure from the final stage (*l* = 4) as the output representation:

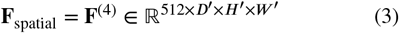

Where *D*^′^, *H*^′^, *W* ^′^ denote the spatial dimensions downsampled by a factor of 16 relative to the input. Consequently, the MRI and PET encoders output their respective spatial feature maps:

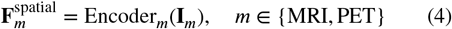

#### Clinical Feature Encoder

Regarding the clinical modality, the data utilized in this study comprises 11-dimensional features, including age, gender, education level, MMSE, ADAS-cog, and marital status. To process this, we designed a dual-branch clinical feature encoder. To simplify the mathematical notation of the Multi-Layer Perceptron (MLP), we first define a fundamental feature mapping operator Φ:

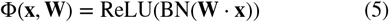

Where · denotes matrix multiplication. Based on this operator, the dual-branch encoder is defined as follows:

Branch I: Disease Stage Embedding. This branch is specifically designed to encode the patient’s disease progression information. It yields a 128-dimensional stage embedding vector, denoted as **e**_stage_, which serves as a global guidance signal:

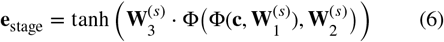

Where **e**_stage_ ∈ ℝ^128^. The network layer dimensions are sequentially 11 → 64 → 128 → 128, with each layer followed by BN and ReLU activation, except for the final layer which uses Tanh activation. The dropout rate is set to 0.35.

Branch II: Clinical Feature Vector. This branch encodes the clinical data into a 512-dimensional feature vector, which is designed for subsequent tri-modal fusion with imaging features during the feature fusion stage:

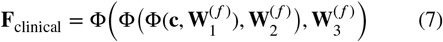

Where **F**_clinical_ ∈ ℝ^512^. The network layer dimensions are sequentially 11 → 64 → 128 → 512, with each layer followed by Batch Normalization and ReLU activation, and the dropout rate is set to 0.35.

The dual-branch design decouples clinical information into two distinct roles. The disease stage embedding, **e**_stage_, encodes high-level stage semantics as a guidance signal, while the clinical feature vector, **F**_clinical_, preserves comprehensive metrics for fusion with imaging features. This separation enables the learning of discriminative guidance signals while retaining complete clinical information for prediction.

### 3.2. Clinical-Guided Spatial Attention

Clinically, imaging manifestations evolve with disease progression: early-stage AD typically presents with localized atrophy (e.g., in the hippocampus) and metabolic deficits, whereas late stages involve widespread cortical damage. Consequently, a robust model must dynamically modulate spatial attention based on the disease stage. Unlike existing approaches that process modalities in isolation, we leverage clinical information as prior knowledge. We propose CGSA to explicitly steer the model’s focus toward spatial regions most relevant to the specific disease stage. The detailed architecture of the CGSA module is illustrated in the top-left panel of Fig. 3.

**Figure 3.**
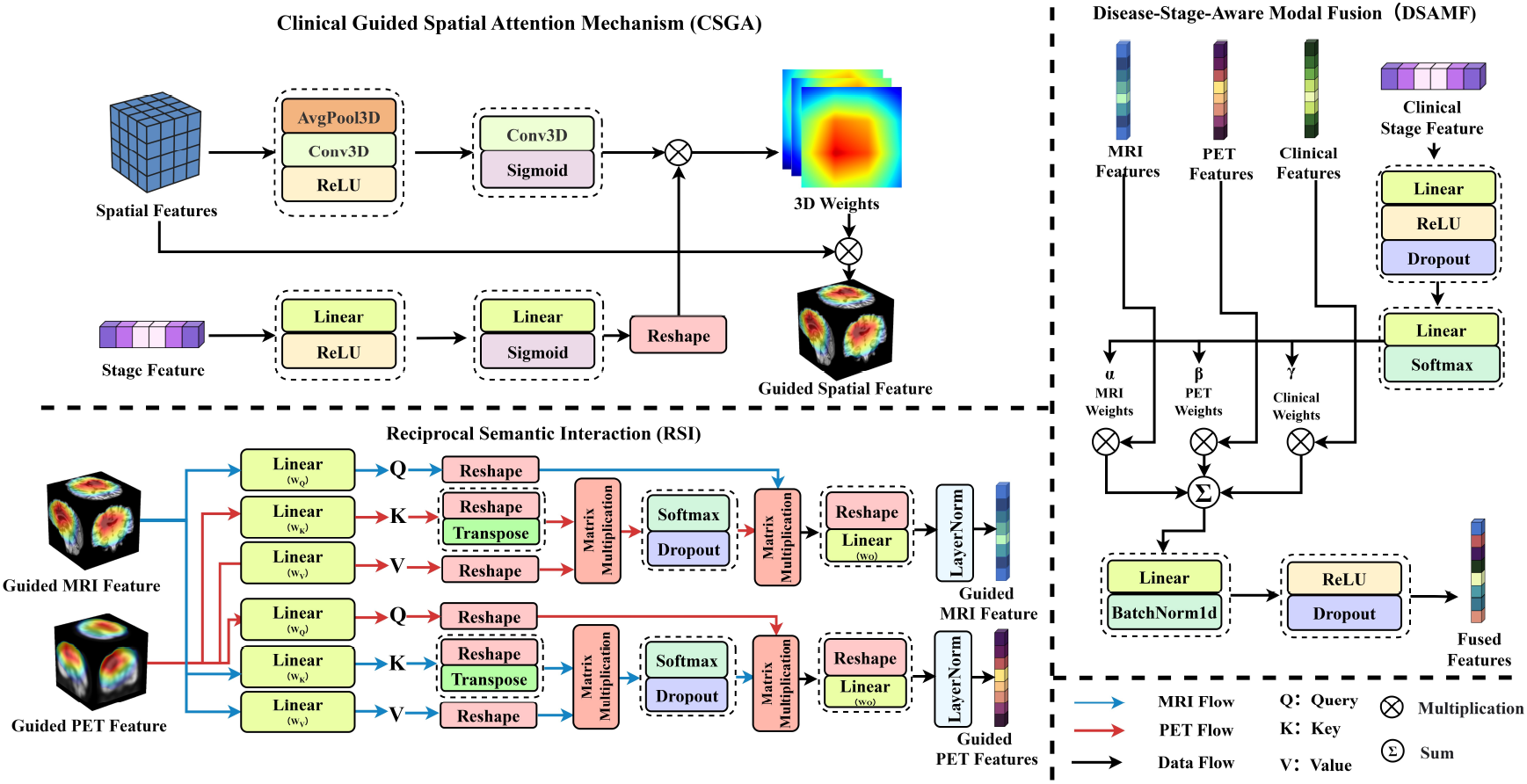
Detailed architecture of the three modules included in COMPASS. The CSGA mechanism modulates spatial features using stage priors, followed by the RSI module for cross-modal feature alignment. The DSAMF module performs dynamic feature integration using learnable importance weights based on the patient’s clinical stage.

Formally, given the spatial feature map **F**_spatial_ extracted by the image encoder and the disease stage embedding **e**_stage_ generated by the clinical encoder, we first map the disease stage embedding to channel attention weights:

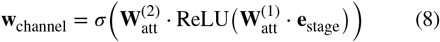

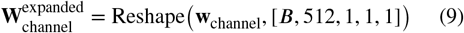

Where 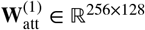 and 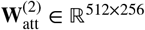 are learnable weight matrices, and *σ*(·) denotes the Sigmoid function. We then expand the dimensions to match the spatial features.

Simultaneously, to capture visual context, we generate spatial-adaptive attention weights by applying GAP followed by channel reduction and expansion operations on the spatial feature map:

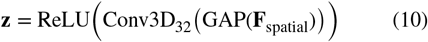

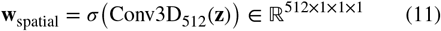

Where the channel dimension is reduced from 512 to 32 and then expanded back to 512. This reduction-expansion strategy facilitates the learning of non-linear relationships between channels.

Subsequently, the clinical-guided channel weights and the spatial-adaptive weights are fused via element-wise multiplication to obtain the combined attention weights:

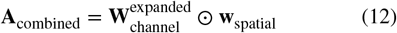

This fusion ensures feature activation only if visual significance coincides with disease relevance, thereby combining clinical prior knowledge with data-driven attention. Finally, the combined weights are applied to the spatial feature map:

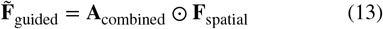

Following global average pooling, we obtain the clinically-guided global feature vector:

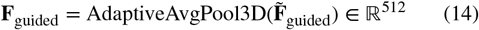

To accommodate the distinct characteristics of MRI and PET modalities, we instantiate independent Clinical-Guided Attention Modules for each. Therefore, separate modules are required to learn modality-specific feature representations:

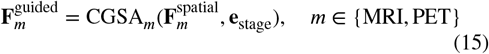

### 3.3. Reciprocal Semantic Interaction

Clinicians routinely combine MRI and PET imaging to improve diagnostic accuracy through comparative analysis. To mimic this, we propose a bidirectional RSI Module aimed at exploiting the complementary nature of these modalities. As depicted in the bottom-left panel of Fig. 3, this module facilitates reciprocal information exchange to bridge the semantic gap between modalities.

We employ Multi-Head Attention [36] to implement cross-modal interaction between 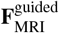 and 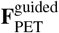. This approach enables parallel attention across different feature subspaces, capturing diverse cross-modal relationships and boosting model robustness.

The mechanism functions by letting one modality query the other. For instance, when MRI queries PET, the MRI feature acts as the Query, while the PET feature serves as both Key and Value. This configuration allows MRI features to adaptively retrieve complementary metabolic information from PET, conditioned on their own structural context. Specifically, we utilize an 8-head attention configuration (head dimension: 64), where projections for Query, Key, and Value are defined as:

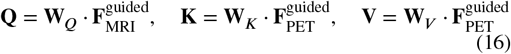

Where **W**_*Q*_, **W**_*K*_, **W**_*V*_ ∈ ℝ^512×512^ denote learnable linear projection matrices. Subsequently, **Q, K**, and **V** are partitioned into *h* = 8 parallel heads, allowing each head to independently process distinct feature subspaces:

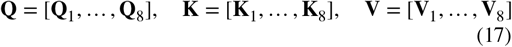

For each head *i* (with dimension *d*_*k*_ = 64), we compute the attention weights of MRI with respect to PET using scaled dot-product attention, followed by a weighted aggregation of the PET features:

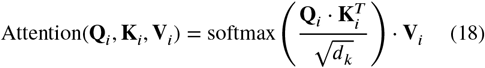

Where the scaling factor 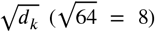 is employed to prevent the dot products from becoming excessively large, which would lead to vanishing gradients in the softmax function. The attention weights generated by softmax 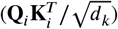 reflect which parts of the PET features the MRI features at distinct positions need to retrieve information from.

Subsequently, the outputs from all heads are concatenated and linearly transformed via an output projection matrix:

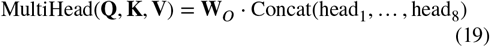

Where **W**_*O*_ ∈ ℝ^512×512^. Finally, employing residual connections and layer normalization, we obtain the MRI-enhanced features fused with PET metabolic information:

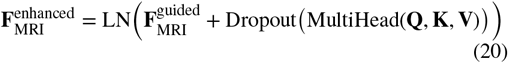

Where LN(·) denotes the Layer Normalization operation, and the dropout rate is set to 0.15. Similarly, we obtain the PET-enhanced features 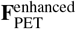 by swapping the query and key-value roles.

To achieve bidirectional interaction, we construct a symmetric mechanism for PET querying MRI, where PET features serve as the Query to retrieve complementary structural context. The PET-enhanced features are formulated as:

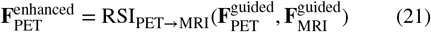

Through this reciprocal process, 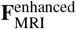 incorporates metabolic cues (e.g., hypometabolism), while 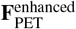 integrates structural details (e.g., atrophy). This dual-enhancement strategy effectively captures intrinsic cross-modal correlations—particularly the structure-metabolism mismatches characteristic of early-stage AD—thereby generating comprehensive representations for the subsequent disease-stage aware fusion.

### 3.4. Disease-Stage-Aware Modal Fusion

Unlike traditional approaches relying on static concatenation or averaging, we propose a DSAMF mechanism to dynamically integrate multi-modal features, as shown in the right panel of Fig. 3. Conventional methods often overlook the heterogeneity of disease progression, whereas clinical reality suggests that the diagnostic value of modalities varies across stages.

To address this, we design a fusion weight generator that adaptively adjusts modal importance based on the patient’s specific condition. As illustrated in the DSAMF module diagram, the weight generation process employs a hierarchical mapping strategy. Specifically, the disease stage embedding **e**_stage_ (from Section 3.1) is first processed by a projection block consisting of a linear layer, ReLU activation, and Dropout to extract robust guidance cues **h**:

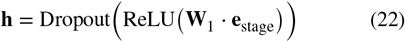

Where **W**_1_ ∈ ℝ^128×128^. This step introduces non-linearity and prevents overfitting. Subsequently, the projected features are mapped to the modal attention scores via a second linear layer and normalized by a Softmax function:

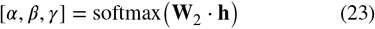

Where **W**_2_ ∈ ℝ^3×128^. The resulting coefficients *α* (MRI), *β* (PET), and *γ* (Clinical) represent the adaptive importance of each modality, satisfying *α* + *β* + *γ* = 1. Utilizing these weights, the fused representation is computed via weighted aggregation:

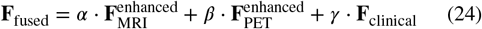

Finally, to model complex feature interactions beyond linear summation, we apply a non-linear transformation block (Linear-BN-ReLU-Dropout) to the fused features:

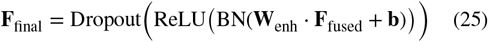

Where **W**_enh_ ∈ ℝ^512×512^. This resulting high-level representation **F**_final_ encapsulates comprehensive multi-modal semantics, serving as the robust input for the downstream prediction tasks.

### 3.5. Multi-Task Disease Classification

Finally, utilizing the fused representation **F**_final_, we design a multi-task classification framework to address three distinct diagnostic levels simultaneously: Task 1 (pMCI vs. sMCI) for early intervention, Task 2 (AD vs. CN) for definitive diagnosis, and Task 3 (AD vs. MCI vs. CN) for comprehensive staging.

First, to extract compact features shared across tasks, **F**_final_ ∈ ℝ^512^ passes through a shared dimensionality reduction block:

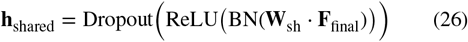

Where **W**_sh_ ∈ ℝ^256×512^ is the weight matrix, and the dropout rate is 0.50. The resulting **h**_shared_ ∈ ℝ^256^ serves as the common input for all downstream tasks.

Subsequently, we instantiate three independent classification heads. To avoid redundancy, we formulate the unified operation for the *k*-th task (*k* ∈ {1, 2, 3}):

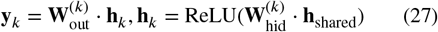

Where 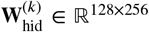 denotes the hidden layer weights. The output weights 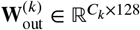 map features to logits, where *C*_*k*_ represents the number of classes (*C*_1_ = *C*_2_ = 2 for binary tasks, *C*_3_ = 3 for the ternary task).

To mitigate class imbalance, we employ Focal Loss [37]. For a given sample with ground truth probability *p*_*t*_, the loss is defined as:

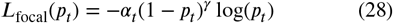

Where *α*_*t*_ balances inverse class frequency, and *γ* = 2.0 is the focusing parameter that down-weights easy examples to prioritize hard-to-classify cases. The total classification loss is the weighted sum of the individual task losses:

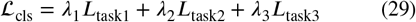

Where *λ*_1_, *λ*_2_, *λ*_3_ are hyperparameters tuned via cross-validation. This multi-task learning paradigm enables shared feature representation, facilitating mutual reinforcement between tasks and enhancing model generalization.

### 3.6. Dual-Head Joint Survival Risk and Time Prediction Network

Conventional classification methods often overlook prognostic questions regarding the precise timing and risk level of disease conversion. To address this, we propose the DH-Net, as illustrated in Fig. 4, which utilizes the frozen feature extraction layers from the previous stage and employs a shared feature extractor followed by a dual-head output mechanism to perform simultaneous time prediction and survival analysis.

**Figure 4.**
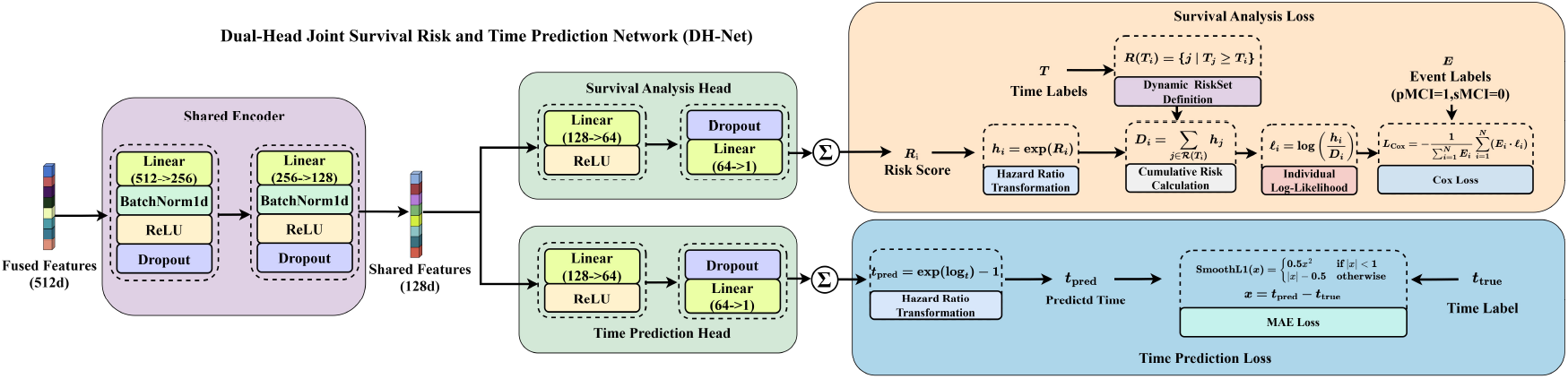
Detailed architecture of the Dual-Head Joint Survival Risk and Time Prediction Network (DH-Net). The network utilizes a Shared Encoder to extract high-level representations from fused features. These are subsequently processed by two parallel branches: the Survival Analysis Head for risk stratification via Cox loss, and the Time Prediction Head for precise conversion time estimation.

Given the high correlation between these two tasks, a shared feature extraction strategy is employed. To accurately yet concisely describe the two-layer stacked architecture (512 → 256 → 128) shown in the architecture diagram, we formulate the propagation recursively using the operator Φ from Section 3.1:

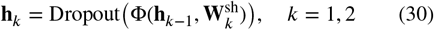

Where we initialize **h**_0_ = **F**_final_, and the final shared representation is **h**_shared_ = **h**_2_ ∈ ℝ^128^. This recursive formulation compactly represents the deep feature extraction process while ensuring rigorous regularization.

Subsequently, independent heads perform time prediction and risk assessment. The time head operates in log-space to compress long-term variances and stabilize gradients. To prevent gradient vanishing, we incorporate a residual connection scaled by a factor of 0.1. The prediction is defined as:

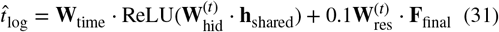

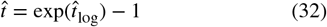

Simultaneously, the risk head outputs a score for patient ranking, where higher scores indicate a greater probability of imminent conversion:

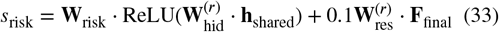

Where **W**_res_ denotes the residual weight matrix used to pass fused information directly to the output. Time prediction is optimized using the Smooth L1 loss [38] to enhance robustness against outliers:

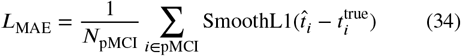

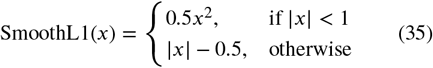

For risk ranking, although sMCI samples lack precise conversion times, they are utilized via the Cox Proportional Hazards model. For each event time *T*_*i*_, we define a dynamic risk set ℛ(*T*_*i*_) = {*j* ∣ *T*_*j*_ ≥ *T*_*i*_}, representing individuals still at risk. The cumulative hazard *D*_*i*_ for this set is computed as:

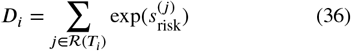

The log-partial likelihood *ℓ*_*i*_ evaluates whether the model correctly assigns higher risk to the subject converting at *T*_*i*_:

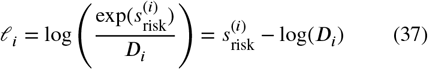

The final Cox loss *L*_Cox_ is the negative average of *ℓ*_*i*_ across all conversion events (*E*_*i*_ = 1), forcing the model to assign higher scores to patients with earlier conversion times:

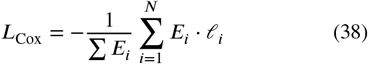

Finally, the model is optimized via a joint loss function to simultaneously address both tasks:

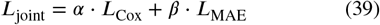

Where *α* = 1.0 and *β* = 0.1 are hyper parameters tailored to balance the relative risk ranking and absolute conversion time prediction, thereby yielding a comprehensive assessment of disease progression.

## 4. Experiments

### 4.1. Dataset

This study was performed on the Alzheimer’s Disease Neuroimaging Initiative (ADNI) dataset [39], including ADNI-1, ADNI-GO, ADNI-2, and ADNI-3. As a landmark North American research initiative, ADNI provides multi-modal AD-related data covering MRI, PET imaging, genetics, cognitive tests, cerebrospinal fluid and blood biomarkers. A total of 1,068 participants with longitudinal follow-up records were recruited, all with paired sMRI, PET, and clinical data. Unlike many methods that adopt dozens of features (e.g., genetic information), we selected 7 readily accessible clinical variables: age, gender, years of education, MMSE score, ADAS-Cog 11 score, marital status, and treatment status. These participants were classified into four groups: CN, pMCI, sMCI, and AD. pMCI and sMCI belong to MCI, distinguished by whether MCI patients converted to AD within 3 years (converters as pMCI, non-converters as sMCI). The sample sizes were 317 CN, 211 pMCI, 356 sMCI, and 184 AD cases, respectively. Detailed demographic characteristics of all participants are summarized in Table 1.

**Table 1.** Demographic and Clinical Characteristics of the Study Population.

| Category | Samples | Male/Female | AGE | Education | MMSE | ADAS | Marry (1/2/3/4/5) |
| --- | --- | --- | --- | --- | --- | --- | --- |
| CN | 317 | 152/165 | 74.08±5.84 | 16.36±2.75 | 29.16±1.03 | 5.78±2.92 | 216/47/33/21/0 |
| PMCI | 211 | 120/91 | 73.53±7.23 | 16.03±2.68 | 27.26±1.92 | 11.83±4.39 | 167/26/13/5/0 |
| SMCI | 356 | 201/155 | 72.08±7.95 | 16.02±2.84 | 28.06±1.76 | 8.16±3.67 | 260/35/45/12/4 |
| AD | 184 | 109/75 | 75.24±7.64 | 15.24±2.90 | 22.99±2.44 | 19.89±7.18 | 162/13/6/3/0 |
| Total | 1068 | 582/486 | 73.51±7.26 | 15.99±2.81 | 27.36±2.74 | 10.20±6.60 | 805/121/97/41/4 |

Note: Marry: 1=Married, 2=Widowed, 3=Divorced, 4=Never married, 5=Unknown.

### 4.2. Evaluation Protocol

For the three AD diagnostic classification tasks (pMCI vs. sMCI, AD vs. CN, and AD vs. MCI vs. CN), we evaluate model performance using standard metrics, including Accuracy (ACC), Area Under the Curve (AUC), Specificity (SPE), Sensitivity (SEN), and F1-score. For the ternary task (AD vs. MCI vs. CN), we employ a One-vs-Rest (OvR) strategy and report macro-averaged results to ensure balanced evaluation across all categories.

For the time prediction (regression) task, evaluation is performed exclusively on pMCI subjects using explicit conversion-time labels. We employ Mean Absolute Error (MAE) and Root Mean Square Error (RMSE) to measure prediction magnitude, alongside time-specific prediction accuracies (Acc@12M, Acc@18M) to assess prognostic precision at key clinical timepoints. The calculation formulas are defined as follows:

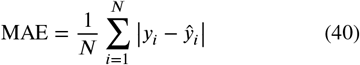

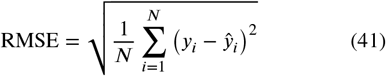

Where *N* denotes the total number of pMCI subjects, and *y*_*i*_ and ŷ_*i*_ represent the ground-truth and predicted conversion times for the *i*-th subject, respectively. Additionally, Acc@*T* measures the percentage of correctly predicted conversions occurring within *T* months (e.g., 12 or 18 months).

For survival analysis, we evaluate the model’s risk assessment capability using three key metrics: Concordance Index (C-index), Time-Dependent AUC (TD-AUC), and Hazard Ratio (HR) with its 95% Confidence Interval (95% CI). Their mathematical definitions are as follows:

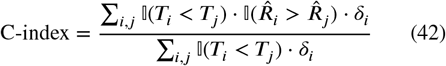

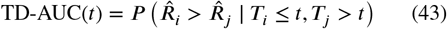

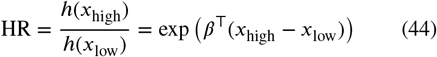

where *T*_*i*_ denotes the observed follow-up time (survival or censoring) for the *i*-th subject, 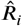 is the predicted risk score, and *δ*_*i*_ is the event indicator (*δ*_*i*_ = 1 indicates conversion, *δ*_*i*_ = 0 indicates censoring). I(·) represents the indicator function. *t* is the specific time horizon. For the Hazard Ratio (HR), *h*(*x*) represents the hazard function of the Cox model, *β* denotes the regression coefficient vector, and *x*_high_ and *x*_low_ represent the feature vectors of the high-risk and low-risk groups, respectively, stratified by the median risk score. An HR > 1 (with a 95% CI lower bound > 1) indicates a significantly elevated risk of disease progression in the high-risk group.

### 4.3. Implementation Details

All paired sMRI and PET images underwent standardized preprocessing. For sMRI, we performed motion correction, intensity normalization, and skull stripping using FreeSurfer [41], followed by registration to the Colin 27 template [42] via the FLIRT tool in FSL [43]. PET images were subjected to spatial normalization, smoothing, and template registration. All brain images were cropped to remove non-brain backgrounds, padded to a cubic shape, and resampled to a uniform resolution of 128×128×128. To enhance model robustness, we applied extensive 3D data augmentation, including random rotation, scaling, and flipping. To simulate anatomical variability, we introduced fast elastic deformation. Additionally, Gaussian noise and random intensity shifts were applied to simulate imaging artifacts and scanner variations. Clinical data were also standardized. The seven features were categorized into numerical (Age, Education, MMSE, ADAS) and categorical (Gender, Marital Status, Treatment). Numerical features were scaled via Z-score normalization, while categorical features were converted to one-hot encodings. All clinical inputs were finally concatenated into a 11-dimensional feature vector.

The COMPASS framework was implemented in Python 3.10 and PyTorch [27], with all experiments conducted on an NVIDIA GeForce RTX 5080 GPU. Following standard protocols [14, 15], we employed 5-fold cross-validation to ensure robust evaluation. The model was trained end-to-end for 200 epochs using the AdamW optimizer [44] (initial learning rate 3 × 10^−5^, weight decay 3 × 10^−5^). We utilized Cosine Annealing with Warm Restarts (*T*_0_ = 30, *T*_*mult*_ = 2, *η*_*min*_ = 1 × 10^−7^) and a linear warmup mechanism [45] for the first 15 epochs to stabilize early training. The batch size was set to 4, and early stopping was implemented with a patience of 50 epochs to prevent overfitting.

### 4.4. Comparison with State-of-the-art Methods

To comprehensively evaluate the effectiveness of the proposed method, we compare it against ten representative baseline methods. To ensure a fair comparison, all experiments were conducted using a unified preprocessing pipeline with identical data resolution (128 × 128 × 128), while data inputs for specific modalities followed the protocols reported in the original studies. These baselines include DA-MIDL [30], which employs dual-attention and multi-instance pooling to identify localized brain atrophy; MPS-FAA [40], which extracts multi-scale features from three MRI planes (sagittal, coronal, axial) using hybrid attention; HOPE [32], which utilizes harmonic graph attention networks to model relational features between brain regions; ADNet9, which disentangles disease-specific pathology from age-related atrophy via feature orthogonal modules; Castellano et al. [46], a 3D CNN architecture that performs AD diagnosis through multimodal feature concatenation; ADFOUND [11], a masked autoencoder framework that leverages both paired and unpaired MRI-PET data; Dia-Mond [12], a ViT-based model that fuses MRI and PET via self-attention and disease-specific augmentations; UNICORS [15], which integrates MRI, PET, and metadata through separate unimodal paths and cross-modal interactions; MultimodalAD [13], a flexible framework using image-tabular Transformers to adapt to varying clinical data combinations; and TriLightNet [14], a lightweight three-branch network using cascaded attention for sMRI, PET, and clinical data interaction.

#### pMCI VS. sMCI

In the highly challenging pMCI vs. sMCI classification task, the proposed COMPASS model demonstrates superior performance, outperforming both unimodal and multimodal baselines. As shown in Table 2, COMPASS achieves the highest accuracy (ACC: 83.19%) and area under the curve (AUC: 83.43%). Compared to recent trimodal methods like TriLightNet (73.46%) and ADFOUND (72.43%), COMPASS yields significant accuracy improvements of 9.73% and 10.76%, respectively. Unlike models that passively extract imaging features, our CGSA module utilizes clinical cognitive embeddings as “prior context” to actively guide the 3D convolutional network toward pathologically significant brain regions. This allows the model to capture subtle discriminative features between the symptomatically similar pMCI and sMCI groups. Furthermore, COMPASS ranks first across key metrics including specificity (SPE: 87.30%), sensitivity (SEN: 76.70%), and F1-score (77.18%). These results confirm that COMPASS effectively integrates MRI, PET, and clinical data to uncover structural and metabolic signals of early conversion, offering a clear advantage for clinical diagnostic support.

**Table 2.**
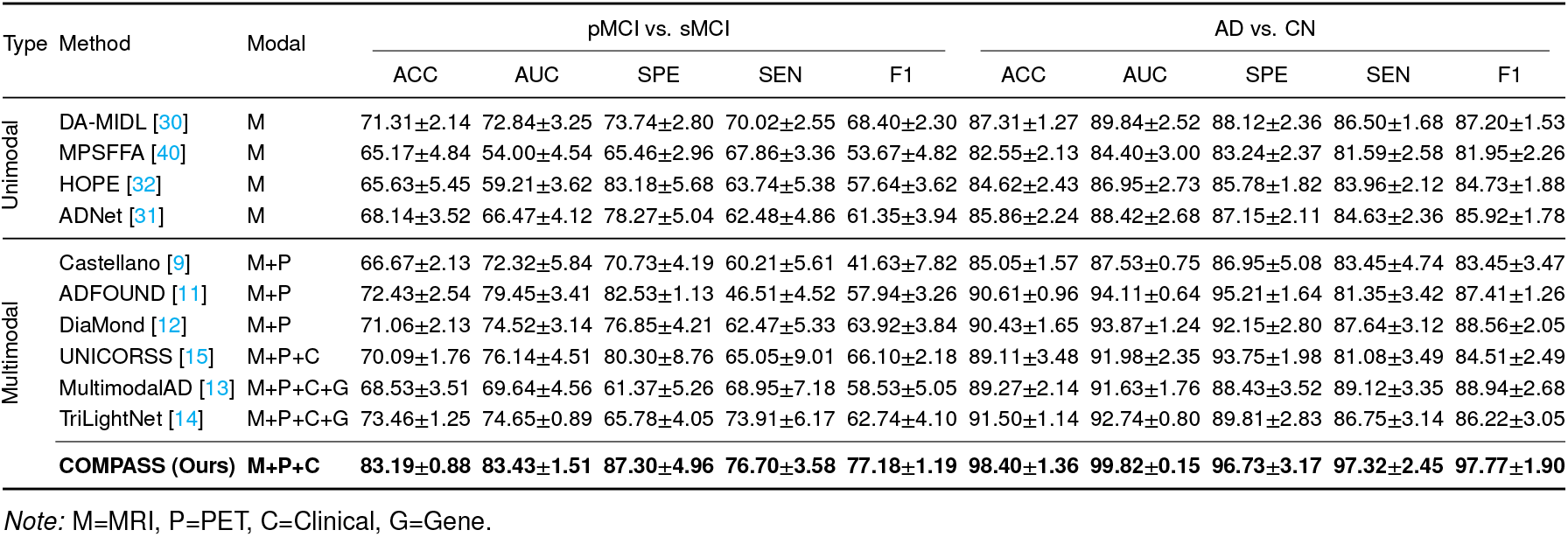
Performance Comparison on pMCI vs. sMCI and AD vs. CN Tasks (Mean ± SD %).

#### AD VS. CN

In the fundamental diagnostic task of AD vs. CN, where patients exhibit pronounced brain atrophy and metabolic abnormalities, model performances are generally higher than in more complex tasks. Nevertheless, COMPASS further pushes the performance boundaries. As shown in Table 2, COMPASS achieves SOTA performance across all key metrics, with an ACC of 98.40% and an AUC of 99.82%. Compared to previously high-performing multimodal methods such as TriLightNet (91.50%) and ADFOUND (79.06%), our approach realizes an absolute accuracy gain of approximately 6.9% to 9.0%. The model’s exceptional balance (SPE: 96.73%, SEN: 97.32%) is primarily attributed to the DSAMF strategy. This mechanism dynamically adjusts the fusion weights of MRI, PET, and clinical data based on disease severity, effectively mimicking the clinical logic where physicians prioritize different biomarkers at various stages. Overall, by deeply integrating multimodal information, COMPASS demonstrates robust feature extraction capabilities, confirming its reliability for definitive diagnosis and establishing a strong performance foundation for comprehensive AD management.

#### AD VS. MCI VS. CN

In the AD vs. MCI vs. CN ternary classification task, the overlap of symptoms during the MCI stage presents a significant challenge. As shown in Table 3, experimental results demonstrate that COMPASS achieves the highest accuracy (ACC: 70.63%) and area under the curve (AUC: 85.03%) in this multi-class challenge. Compared to the previously top-performing multimodal method, TriLightNet (65.68%), COMPASS yields a 4.95% absolute improvement in accuracy and reaches an F1-score of 71.71%, significantly outperforming all other baselines. These findings suggest that the COMPASS framework, through clinical-prior guidance and deep multimodal fusion, effectively captures the heterogeneous variations across disease stages, providing comprehensive coverage of the entire Alzheimer’s progression spectrum.

**Table 3.**
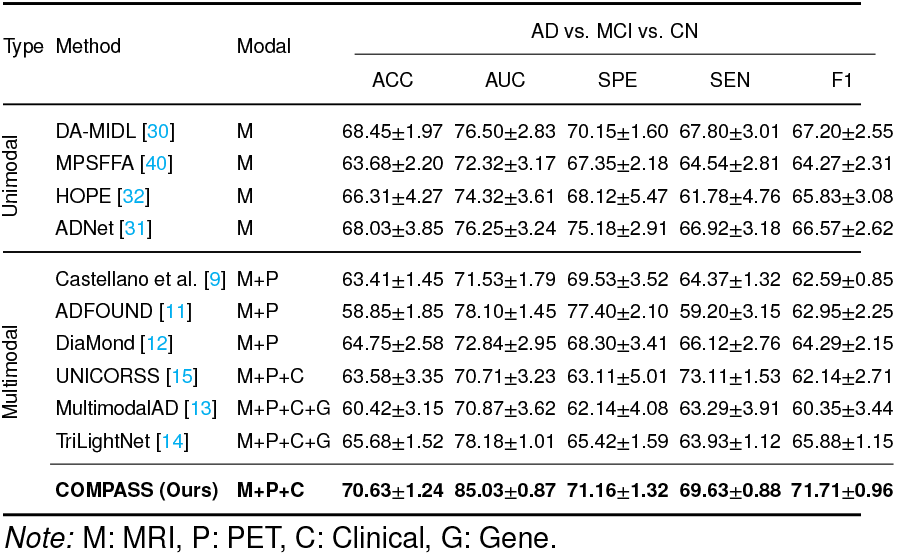
Performance Comparison on AD vs. MCI vs. CN Ternary Task (Mean ± SD %).

Finally, Fig. 5 provides a visual corroboration of these quantitative results. Fig. 5(a) displays the bar charts of key metrics across the three tasks, clearly showing that COMPASS consistently outperforms competing methods in terms of overall effectiveness. Complementing this, Fig. 5(b) illustrates the accuracy distributions from the 5-fold cross-validation using violin plots. In contrast to the wider distributions observed in baseline methods, COMPASS exhibits a more compact and higher-positioned distribution, confirming not only its superior accuracy but also its enhanced stability and robustness against data variations.

**Figure 5.**
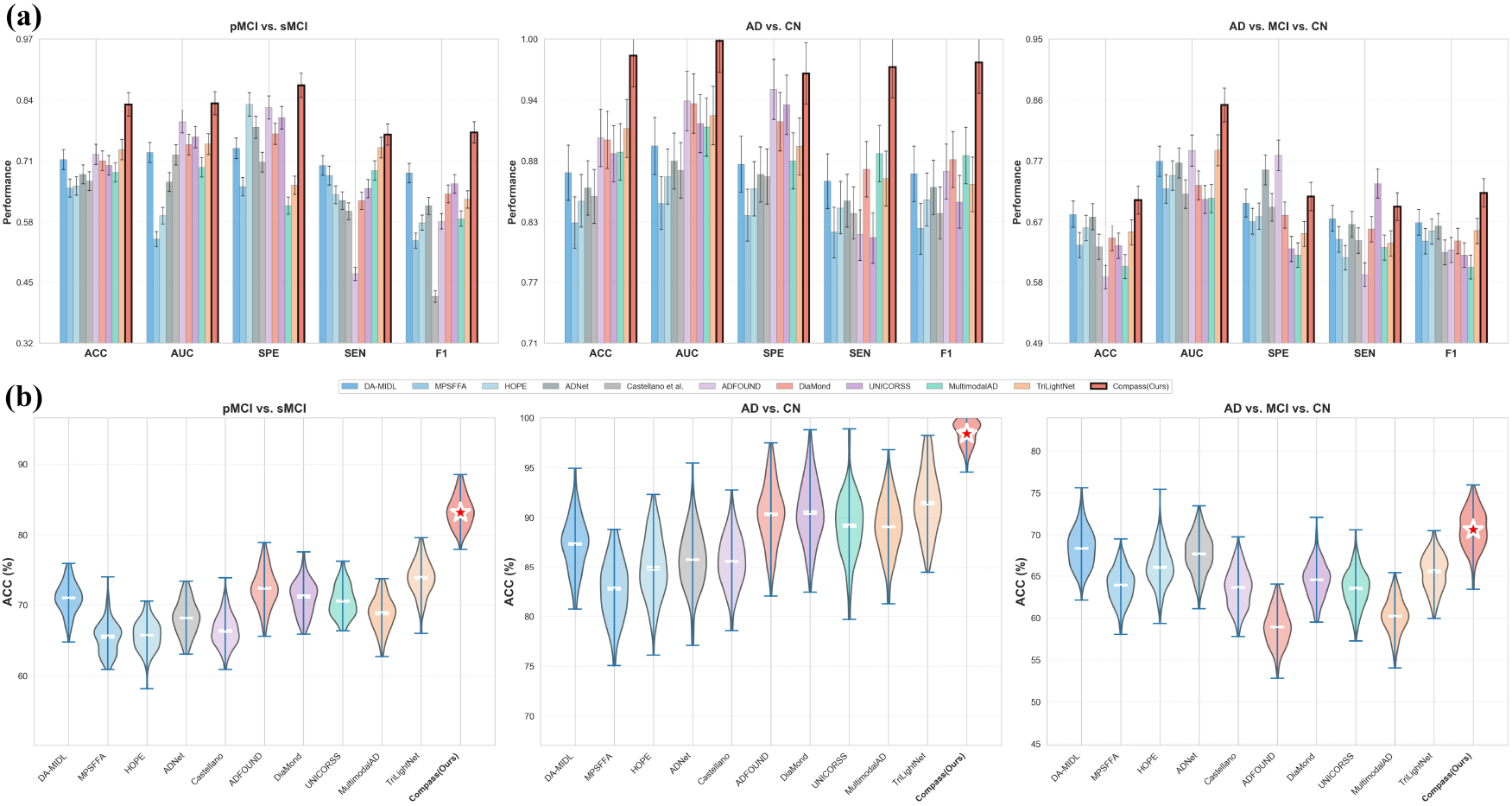
Performance comparison with SOTA methods across diagnostic tasks. (a) Bar charts of key metrics showing COMPASS (red bars) consistently outperforms baselines. (b) Violin plots of accuracy distributions demonstrating the superior stability and robustness of COMPASS (red star).

### 4.5. Conversion Time Prediction and Survival Analysis Testing

Given the continuous and chronic nature of AD, binary classification alone falls short of satisfying clinical requirements for precise risk stratification and intervention timing. Therefore, predicting conversion timing and assessing progression risk (via survival analysis) for MCI patients is of paramount clinical value. To address this, we developed DH-Net. It is noteworthy that most existing SOTA methods, such as ADFOUND and UNICORSS, do not natively support these prognostic functions. To ensure a fair comparison, we adapted these methods by integrating their feature extractors with our DH-Net architecture, and the detailed performance results are presented in Table 4. Thanks to its plug-and-play shared-feature design, DH-Net effectively consolidates fused features from various models and translates them into actionable prognostic information.

**Table 4.** Performance Comparison on Conversion Time Prediction and Survival Analysis Tasks.

| Type | Method | Modal | Conversion Time Prediction |  |  |  | Survival Analysis |  |  |
| --- | --- | --- | --- | --- | --- | --- | --- | --- | --- |
|  |  |  | MAE | RMSE | Acc@12M | Acc@18M | C-index | TD-AUC | HR (95% CI) |
| Unimodal | DA-MIDL [30] | M | 9.89±0.82 | 12.50±1.10 | 68.4%±4.2% | 76.5%±4.1% | 0.742±0.021 | 0.751±0.030 | 2.36 (1.52–3.18) |
|  | MPSFFA [40] | M | 11.92±1.42 | 15.10±1.85 | 60.5%±5.8% | 66.1%±5.2% | 0.668±0.035 | 0.683±0.041 | 1.83 (1.10–2.63) |
|  | HOPE [32] | M | 10.71±1.06 | 13.82±1.54 | 64.8%±4.1% | 71.3%±4.4% | 0.702±0.022 | 0.726±0.025 | 2.11 (1.17–2.64) |
| Multimodal | Castellano [9] | M+P | 10.62±1.15 | 13.92±1.55 | 63.1%±4.8% | 69.4%±5.1% | 0.695±0.021 | 0.715±0.033 | 2.12 (1.35–2.89) |
|  | ADFOUND [11] | M+P | 9.35±0.88 | 11.85±0.94 | 70.2%±3.5% | 78.1%±3.2% | 0.762±0.020 | 0.773±0.025 | 2.65 (1.75–3.55) |
|  | UNICORSS [15] | M+P+C | 8.85±0.76 | 11.30±0.85 | 72.5%±2.9% | 81.6%±3.0% | 0.784±0.018 | 0.798±0.022 | 2.98 (1.95–4.01) |
|  | **COMPASS (Ours)** | **M+P+C** | **7.96±0.68** | **10.73±0.69** | **74.9%±3.1%** | **87.2%±3.4%** | **0.819±0.014** | **0.832±0.044** | **3.42 (2.26–5.17)** |

Note: M=MRI, P=PET, C=Clinical.

#### Conversion Time Prediction

The COMPASS model demonstrates a significant advantage in predicting conversion timing, achieving an MAE of 7.96 ± 0.68 months and an RMSE of 10.73 ± 0.69 months. These results are markedly superior to adapted versions of UNICORSS (MAE: 8.85) and ADFOUND (MAE: 9.35) when integrated with the same DH-Net architecture. Furthermore, COMPASS achieved accuracy scores of 74.9% and 87.2% at the 12-month and 18-month prediction horizons, respectively. This performance not only validates the architectural strengths of DH-Net for temporal regression but also reveals that the features extracted by the COMPASS front-end—via clinical guidance and dynamic fusion—are highly compatible with the DH-Net joint training mechanism. This synergy allows the model to precisely identify the temporal windows of disease conversion.

#### Survival Analysis and Risk Assessment

As a critical component for prognostic assessment within the COMPASS framework, DH-Net also performs survival analysis to achieve effective patient risk stratification. Experimental results further validate the robustness of this module in risk ranking. Regarding the C-index, which measures the discriminative power of the predictive model, COMPASS achieved 81.93 ± 1.36, significantly outperforming unimodal methods (e.g., DA-MIDL: 74.20) and other adapted multi-modal frameworks (e.g., UNICORSS: 78.40). These results indicate that DH-Net effectively leverages the rich semantic features provided by COMPASS to more accurately rank conversion risks across patients. Furthermore, the TD-AUC reached 83.20 ± 4.37, quantifying the model’s discriminative stability throughout the entire observation period. Crucially, HR analysis revealed that COMPASS achieved a value of 3.42 (95% CI: 2.26-5.17), the highest among all comparative experiments. This indicates that through the joint optimization of DH-Net, COMPASS successfully constructs a feature space capable of significantly distinguishing high-risk conversion groups from low-risk stable groups. These findings demonstrate the comprehensive clinical value of the framework in integrating diagnosis, time prediction, and risk assessment.

Finally, we selected two subjects from the validation set to visualize the model’s performance in conversion-time prediction and survival analysis (Fig. 6), demonstrating its precision in individual risk stratification and pathological progression tracking.

**Figure 6.**
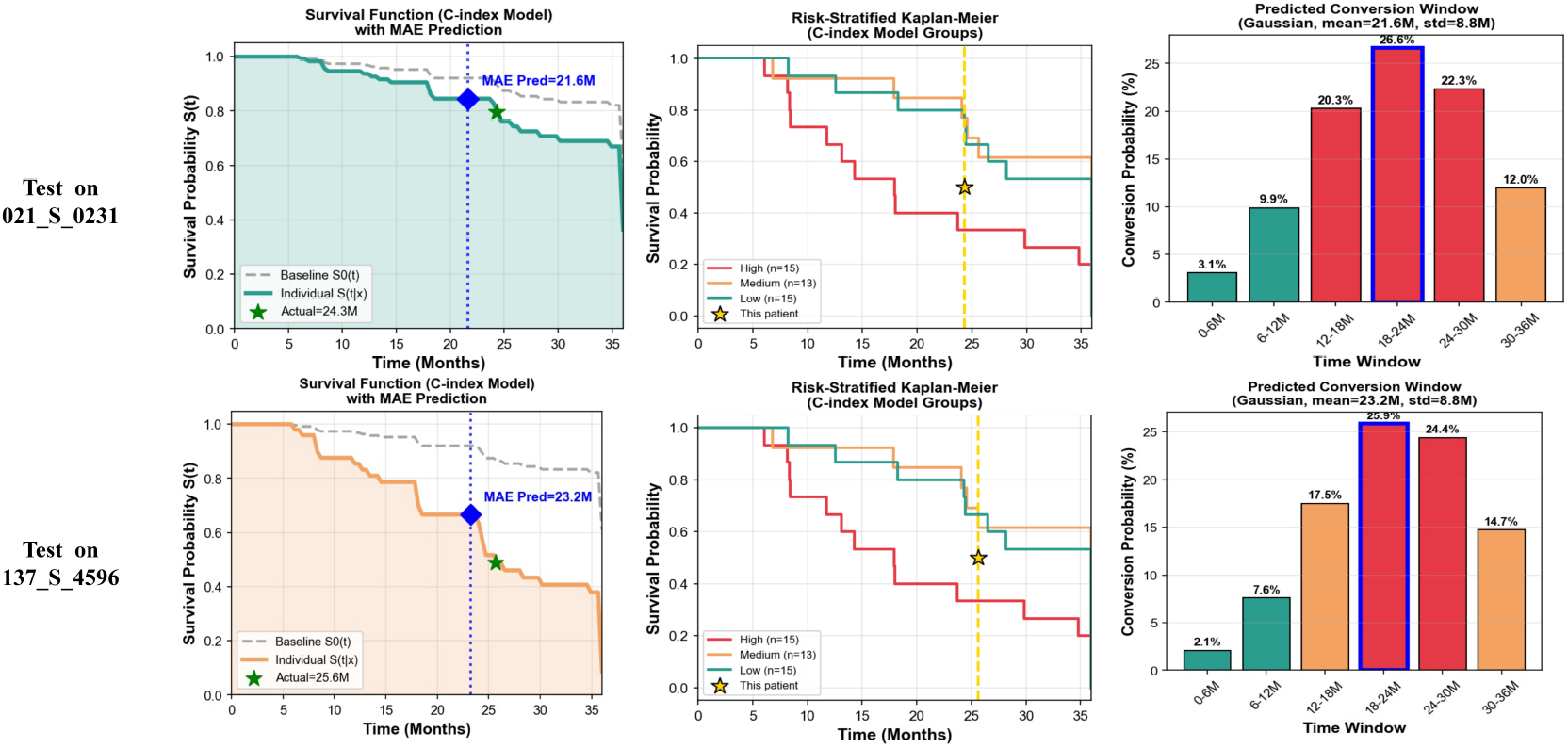
Individualized prognostic visualization for pMCI subjects. The left column displays survival curves (*S*(*t*)), highlighting the alignment between predicted (blue diamond) and actual (green star) conversion times. The middle column illustrates risk stratification against population baselines, while the right column presents the probability distribution of predicted conversion windows.

### 4.6. Ablation Study on Multi-task Synergy

We conducted ablation studies to validate the synergy between the diagnostic tasks (Tasks 1–3) and conversion prediction, comparing independent sub-task training against full-task joint training to investigate shared feature learning effects. Detailed results for the classification tasks are presented in Table 5. Joint training consistently outperforms independent training across all tasks. Notably, in the challenging pMCI vs. sMCI task, accuracy improved from 80.53% to 83.19%. This suggests that multi-task learning fosters generalizable features, where clear pathological patterns from the AD vs. CN task assist in capturing subtle conversion signals during the MCI stage via cross-task knowledge transfer.

**Table 5.** Ablation Study of Classification Performance: Single-task vs. Joint Training.

| Method | Task 1 (AD vs. CN) |  |  | Task 2 (pMCI vs. sMCI) |  |  | Task 3 (AD vs. MCI vs. CN) |  |  |
| --- | --- | --- | --- | --- | --- | --- | --- | --- | --- |
|  | ACC | AUC | F1 | ACC | AUC | F1 | ACC | AUC | F1 |
| Only Task 1 | 80.53±1.13 | 82.73±2.53 | 73.17±1.53 | – | – | – | – | – | – |
| Only Task 2 | – | – | – | 98.21±1.82 | 97.96±0.05 | 97.62±2.33 | – | – | – |
| Only Task 3 | – | – | – | – | – | – | 67.39±1.61 | 76.23±2.11 | 65.59±1.83 |
| **Full** | **83.19±0.88** | **83.43±1.51** | **77.18±1.19** | **98.40±1.36** | **99.82±0.15** | **97.77±1.90** | **70.63±1.24** | **85.03±0.87** | **71.71±0.96** |

The synergy between diagnostic and prognostic tasks is further quantified in Table 6. Compared to single-task regression, joint optimization reduced the conversion time prediction MAE from 11.52 to 7.96 months. Simultaneously, the C-index rose from 71.19 to 81.93, and the HR increased from 1.29 to 3.42. This confirms that DH-Net benefits from the discriminative feature space constructed by classification tasks, enabling the model to characterize not just the disease state but also its severity and temporal progression.

**Table 6.**
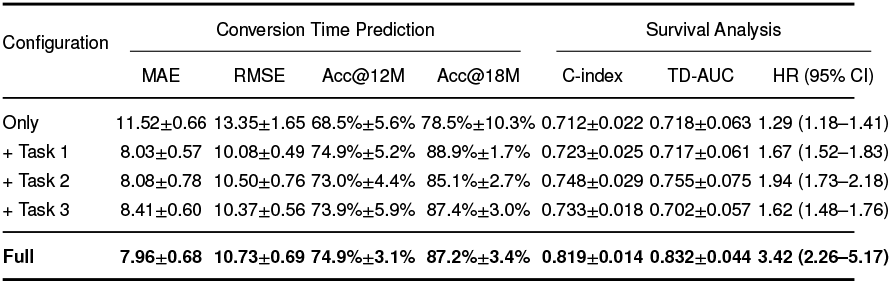
Impact of Multi-task Synergy on Conversion Time Prediction and Survival Analysis.

In summary, the sub-tasks in COMPASS are not isolated but achieve feature complementarity through shared layers and joint loss constraints. This collaborative strategy delivers high-precision diagnosis alongside accurate long-term prognosis, highlighting the effectiveness of multi-task learning for comprehensive AD management.

### 4.7. Ablation Study of Core Components

To investigate the marginal contributions of each core component, we conducted a hierarchical ablation study on the pMCI vs. sMCI task. We established a Baseline comprising basic encoders with feature concatenation and incrementally integrated CGSA, RSI, and DSAMF to elucidate how the framework overcomes performance bottlenecks. Detailed experimental results are presented in Table 7 and visually summarized in the left panel of Fig. 7. Specific results indicate that the Baseline, lacking prior guidance, performed poorly on overlapping pMCI/sMCI representations (ACC: 72.89%, AUC: 73.45%), confirming the susceptibility of traditional CNNs to non-pathological noise. The integration of CGSA yielded the first significant leap (ACC: 75.78%, SEN: 75.43%) by shifting the focus to key anatomical subspaces like the hippocampus. Subsequently, the RSI module bridged the semantic gap between MRI and PET manifolds via cross-modal attention, consistently improving the AUC. At the decision fusion level, the DSAMF module addressed pathological heterogeneity through nonlinear dynamic weighting, independently boosting specificity (SPE) from 70.76% to 74.81%.

**Table 7.**
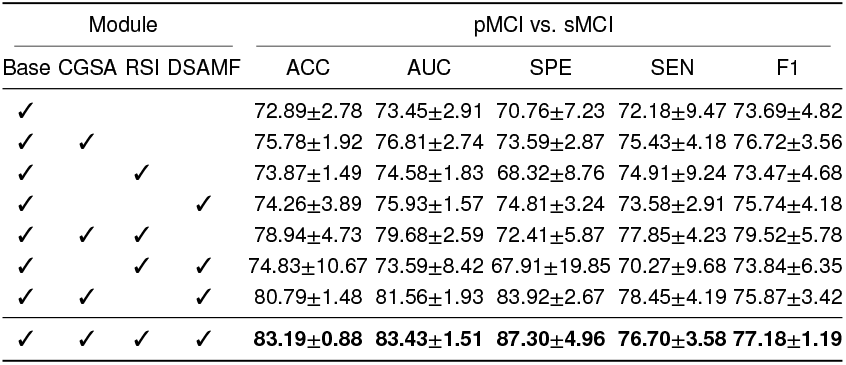
Ablation study of core components.

**Figure 7.**
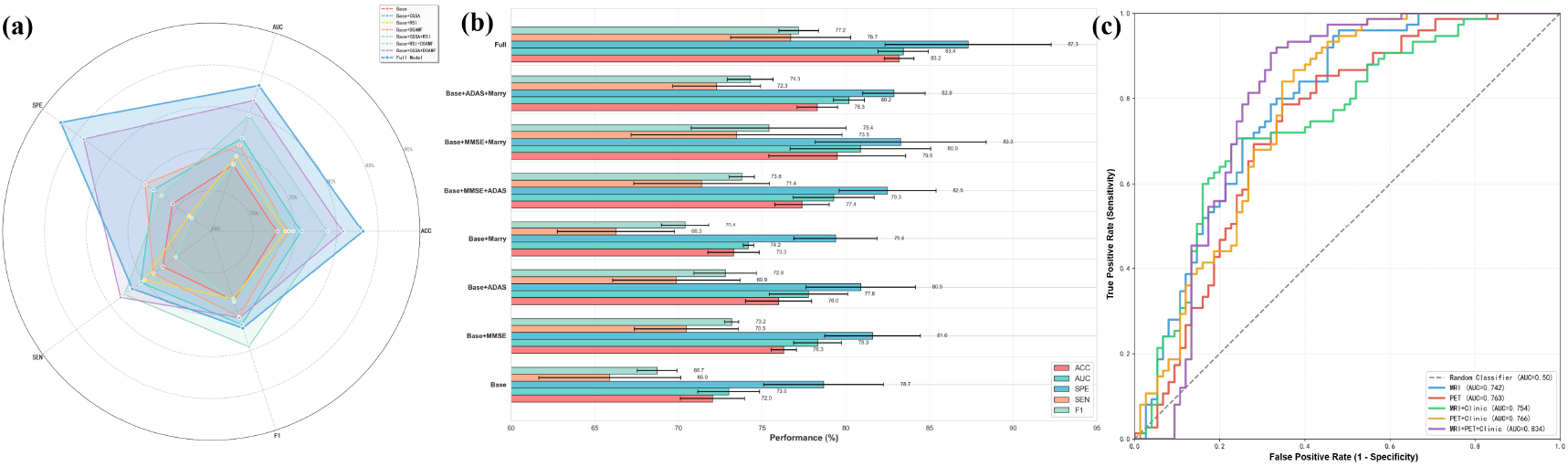
Comprehensive ablation studies of the COMPASS framework. (a) Radar chart exhibiting the impact of key network modules on diagnostic performance. (b) Evaluation of different clinical prior combinations to validate the efficacy of the clinical guidance strategy. (c) ROC curves comparing single-modal and multi-modal inputs, highlighting the superiority of the full fusion framework.

Crucially, the synergistic operation of spatial localization, semantic interaction, and dynamic fusion induces a significant systemic emergence effect. The full COMPASS framework demonstrates a substantial performance lead across all metrics, with ACC increasing to 83.19%, AUC reaching 83.43%, and F1 reaching 77.18%, representing a remarkable 10.30% improvement over the Baseline. These results validate that the framework successfully establishes a closed-loop feature evolution path, progressing from precise localization through semantic synergy, and culminating in dynamic integration. This tripartite design effectively transforms multi-source heterogeneous data into prognostic representations that align closely with clinical diagnostic logic, confirming its theoretical depth and potential for precision medical diagnosis.

### 4.8. Analysis of Different Clinical Data

To quantify the marginal contributions of multi-dimensional clinical priors, we conducted a hierarchical ablation study on the pMCI vs. sMCI task. Detailed experimental results are presented in Table 8 and visually displayed in the middle panel of Fig. 7. Results reveal the limitations of basic demographics; relying solely on age, gender, and education (Base) yielded only 72.03% accuracy, indicating that macroscopic risk factors fail to capture subtle pathological heterogeneity between MCI subtypes.

**Table 8.**
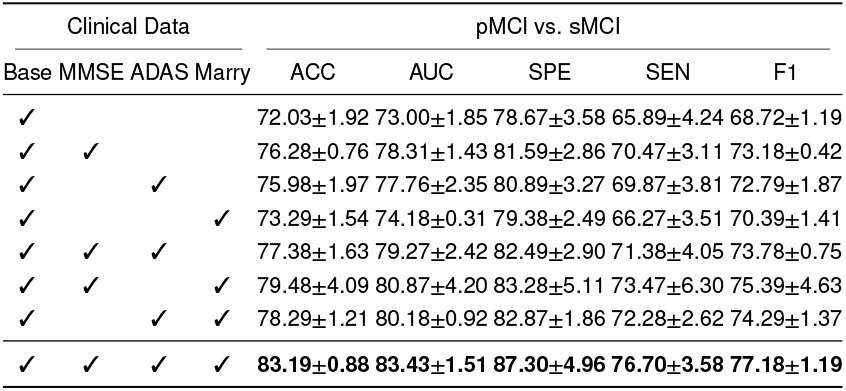
Ablation study on the importance of different clinical data.

Incorporating neuropsychological scales yielded the first significant performance leap. Specifically, adding MMSE alone raised accuracy to 76.28% (AUC 78.31%). This establishes cognitive scales as core phenotypic markers, proving that quantifying memory and executive impairment is essential for distinguishing disease progression. Combinatorial analysis highlighted that orthogonal complementarity outweighs homogeneous superposition. Notably, adding social factors (marital status) to the Base + MMSE model achieved 79.48% accuracy, surpassing the addition of a second cognitive scale (ADAS-cog, 77.38%). This suggests social support factors provide unique information dimensions independent of cognitive scores, addressing blind spots in biomedical indices, whereas dual cognitive scales suffer from redundancy.

Ultimately, the full-dimensional fusion model (Base + MMSE + ADAS + Marital) achieved a peak accuracy of 83.19%, an 11.16% improvement over the Baseline. These results demonstrate that constructing a panoramic clinical portrait—encompassing biological, psychological, and sociological factors—is a prerequisite for overcoming performance bottlenecks in early AD conversion prediction.

### 4.9. Ablation Study of Different Modalities

In this section, we performed a multimodal ablation study using the pMCI vs. sMCI differentiation task as a benchmark to quantify the independent contributions and limitations of different data modalities in disease representation. Detailed experimental results are presented in Table 9 and visually illustrated in the right panel of Fig. 7. It is important to note that since clinical data serves as guidance information fused with imaging data, our experimental configurations do not include a standalone clinical data baseline. Experimental data revealed distinct bottlenecks in single imaging modalities. Sole reliance on structural MRI yielded high sensitivity (81.57%) but poor specificity (63.59%) and accuracy (67.67%), highlighting the high false-positive risk when using anatomical atrophy to exclude non-specific interference such as normal aging. While PET improved accuracy to 71.95%, it remained insufficient to establish a robust decision boundary.

**Table 9.** Ablation study of different modalities.

| Modality |  |  | pMCI vs. sMCI |  |  |  |  |
| --- | --- | --- | --- | --- | --- | --- | --- |
| MRI | PET | Clinic | ACC | AUC | SPE | SEN | F1 |
| ✓ |  |  | 67.67±6.28 | 74.18±4.11 | 63.59±8.69 | 81.57±7.66 | 69.06±2.46 |
|  | ✓ |  | 71.95±5.81 | 76.34±5.13 | 71.04±9.25 | 73.51±9.95 | 66.09±5.99 |
| ✓ |  | ✓ | 69.87±8.06 | 75.37±9.79 | 65.88±9.84 | 78.27±9.51 | 70.11±6.83 |
|  | ✓ | ✓ | 74.02±10.56 | 79.56±5.87 | 73.40±8.11 | 78.24±10.19 | 74.65±7.60 |
| ✓ | ✓ | ✓ | **83.19±0.88** | **83.43±1.51** | **87.30±4.96** | **76.70±3.58** | **77.18±1.19** |

Integrating clinical data provided a critical corrective effect. Adding demographics and cognitive scores (e.g., PET + Clinic) increased accuracy to 74.02%, demonstrating that clinical priors impose semantic constraints that effectively compensate for the specificity deficits of pure imaging analysis. Optimal performance was achieved via full-dimensional fusion (MRI + PET + Clinical). This approach elevated the AUC to 83.43% and accuracy to 83.19%, representing an approximate 9% improvement over the best bimodal combination. Detailed analysis shows that full fusion not only retained high sensitivity (83.19%) but also significantly boosted specificity to 77.18% by integrating PET metabolic information and clinical functional status, achieving an optimal F1 score (83.43%). This validates that heterogeneous data complementarity constructs a panoramic disease portrait, overcoming single-modality blind spots to maximize predictive robustness.

### 4.10. Interpretability Analysis

While the COMPASS framework demonstrates superior quantitative performance across multi-class tasks, model transparency and trustworthiness remain paramount for clinical application. Deep learning models are frequently characterized as black boxes, where a lack of intuitive insight into their decision-making rationale hinders their deployment in computer-aided diagnosis. Consequently, this section employs multi-level interpretability analysis to demystify this black box and unveil the most discriminative features captured by the model. Beyond validating the alignment between the model’s focus regions and established Alzheimer’s neuropathology, we attempt to mine potential neural compensatory mechanisms from the learned weights, providing biological insights into disease progression. We conduct this analysis using the persistent challenge of pMCI vs. sMCI differentiation as a case study.

To elucidate the model’s decision focus when distinguishing pMCI from sMCI, we extracted and visualized the most discriminative brain functional connectivity patterns. As illustrated in Fig. 8(a), 3D brain views and chord diagrams intuitively depict the distribution of connections assigned high weights. Results indicate that these discriminative connections are not diffusely distributed across the whole brain but are heavily anchored within the limbic system, particularly involving neural circuits associated with the hippocampus, parahippocampal gyrus, and temporal lobes. Furthermore, long-range connections involving the precuneus and posterior cingulate cortex (PCC) exhibited significant weights, aligning with the pathological mechanism of functional decoupling within the Default Mode Network (DMN) characteristic of early AD.

**Figure 8.**
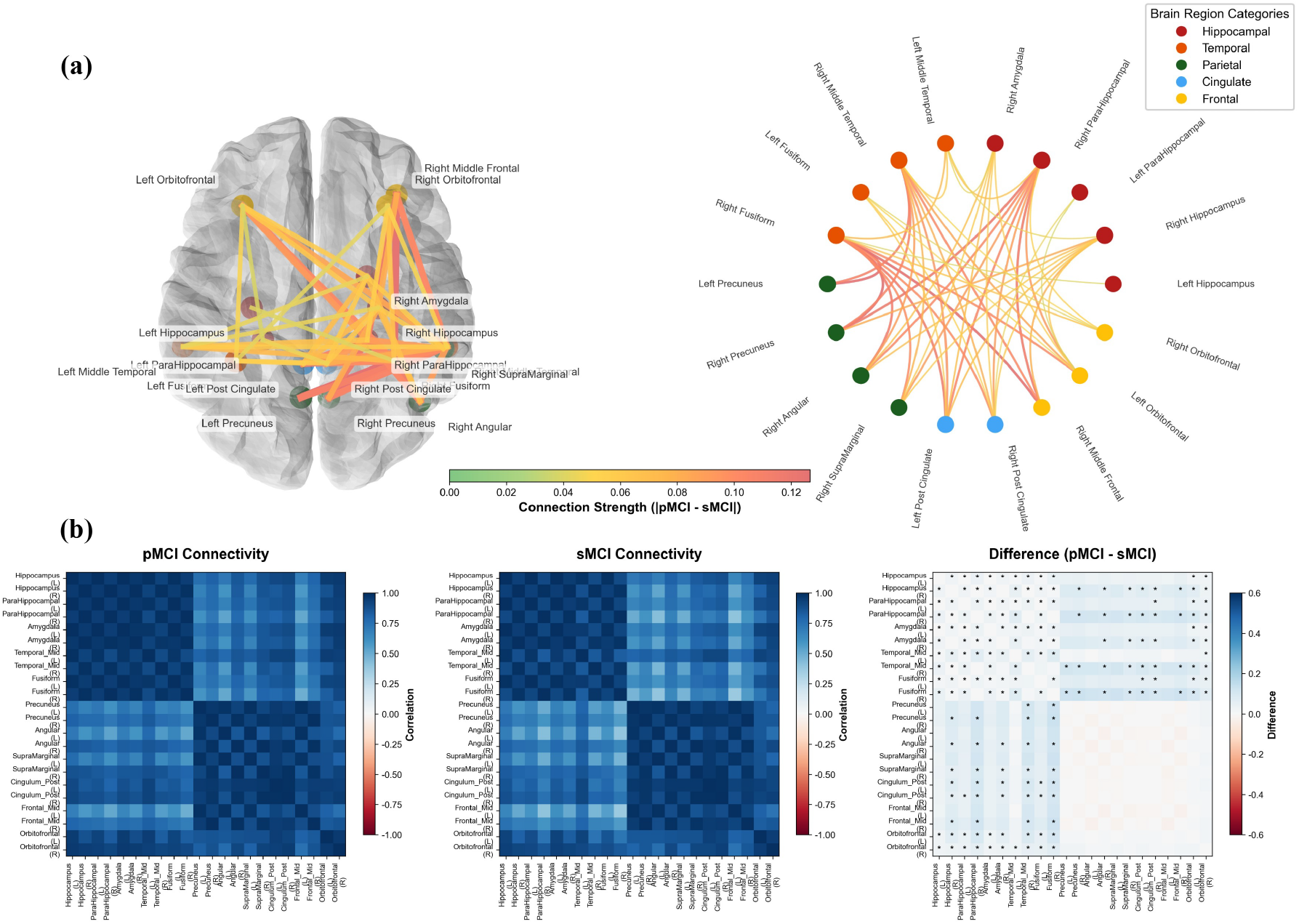
Interpretability analysis of the COMPASS framework. (a) 3D visualizations and chord diagrams reveal that the most discriminative connections are heavily anchored within the limbic system and DMN. (b) Statistical validation via connectivity matrices shows that significant group differences (*p* < 0.05) are clustered in the medial temporal lobe while sparing the frontal cortex, consistent with the trajectory of early AD pathology.

To validate the statistical robustness of these features, we further analyzed the inter-group connectivity matrices and their differential significance, as shown in Fig. 8(b). The differential matrix analysis clearly reveals a spatial heterogeneity characterized by temporal significance with frontal sparing: statistical significance markers (*p* < 0.05) are densely clustered in the top-left region of the matrix, corresponding to the medial temporal lobe system, which encompasses the hippocampus, parahippocampal gyrus, and amygdala. Conversely, the bottom-right region, representing intra-connectivity within the frontal lobe and sensorimotor cortex, shows no significant inter-group differences. This finding aligns precisely with the classical Braak pathological staging trajectory of AD, wherein neurodegeneration initiates in the medial temporal lobe and has not yet severely encroached upon the frontal neocortex during the MCI stage. This provides compelling evidence that the model has successfully locked onto core early-stage lesions rather than relying on diffuse whole-brain background noise for prediction.

### 4.11. Clinical and Pathological Mechanism Analysis

Building upon the preceding interpretability analysis, we further investigated the quantitative importance of individual brain regions to derive deeper neurobiological insights. This analysis transcends validating the model’s attentional foci; rather, it represents a critical re-examination of the pathological mechanisms underlying early AD.

Systematic statistical analysis of attention weights across 22 key brain regions (Table 10) reveals significant hemispheric asymmetry within the medial temporal lobe (MTL) during pMCI vs. sMCI differentiation, characterized by distinct right-hemisphere dominance (Fig. 10). Specifically, the right hippocampus exhibited a significantly higher effect size (Cohen’s *d* = 0.105) and importance ranking (Rank = 6) compared to the left (*d* = 0.055, Rank = 14). This trend was consistently validated in the parahippocampal gyrus and amygdala. These findings align with established hippocampal lateralization, where the right side governs spatial memory and the left handles verbal memory. Our results suggest spatial memory functions are more vulnerable in early AD, corroborating clinical observations of spatial disorientation and implying that right hippocampal atrophy may serve as a more sensitive biomarker than bilateral measurements.

**Table 10.** Hemispheric asymmetry of the hippocampal system (Atten-tion Weight).

| Region | pMCI Attention |  | sMCI Attention |  | Statistics |  |
| --- | --- | --- | --- | --- | --- | --- |
|  | Left | Right | Left | Right | Diff. | p-value |
| Hippocampus | $0.653 \pm 0.089$ | $0.755 \pm 0.092$ | $0.650 \pm 0.095$ | $0.746 \pm 0.098$ | 0.0035 | 0.041 |
| Parahippocampal gyrus | $0.618 \pm 0.076$ | $0.726 \pm 0.084$ | $0.615 \pm 0.081$ | $0.717 \pm 0.087$ | 0.0035 | 0.038 |
| Amygdala | $0.647 \pm 0.082$ | $0.726 \pm 0.088$ | $0.644 \pm 0.086$ | $0.718 \pm 0.091$ | 0.0028 | 0.049 |
| Average | 0.639 | 0.736 | 0.636 | 0.727 | 0.0032 | < 0.05 |

To validate these findings, we visualized high-resolution attention maps (Fig. 9) by projecting feature-layer attention onto anatomical MRI scans. Quantitative observation confirmed right-hemisphere dominance: for both pMCI and sMCI, attention weights on the right hippocampus consistently exceeded the left (e.g., pMCI: *R* = 0.714 vs. *L* = 0.387). Furthermore, pMCI patients exhibited broader and more intense activation regions, indicating the model assigns stronger discriminative weights to pathological features in high-risk converters.

**Figure 9.**
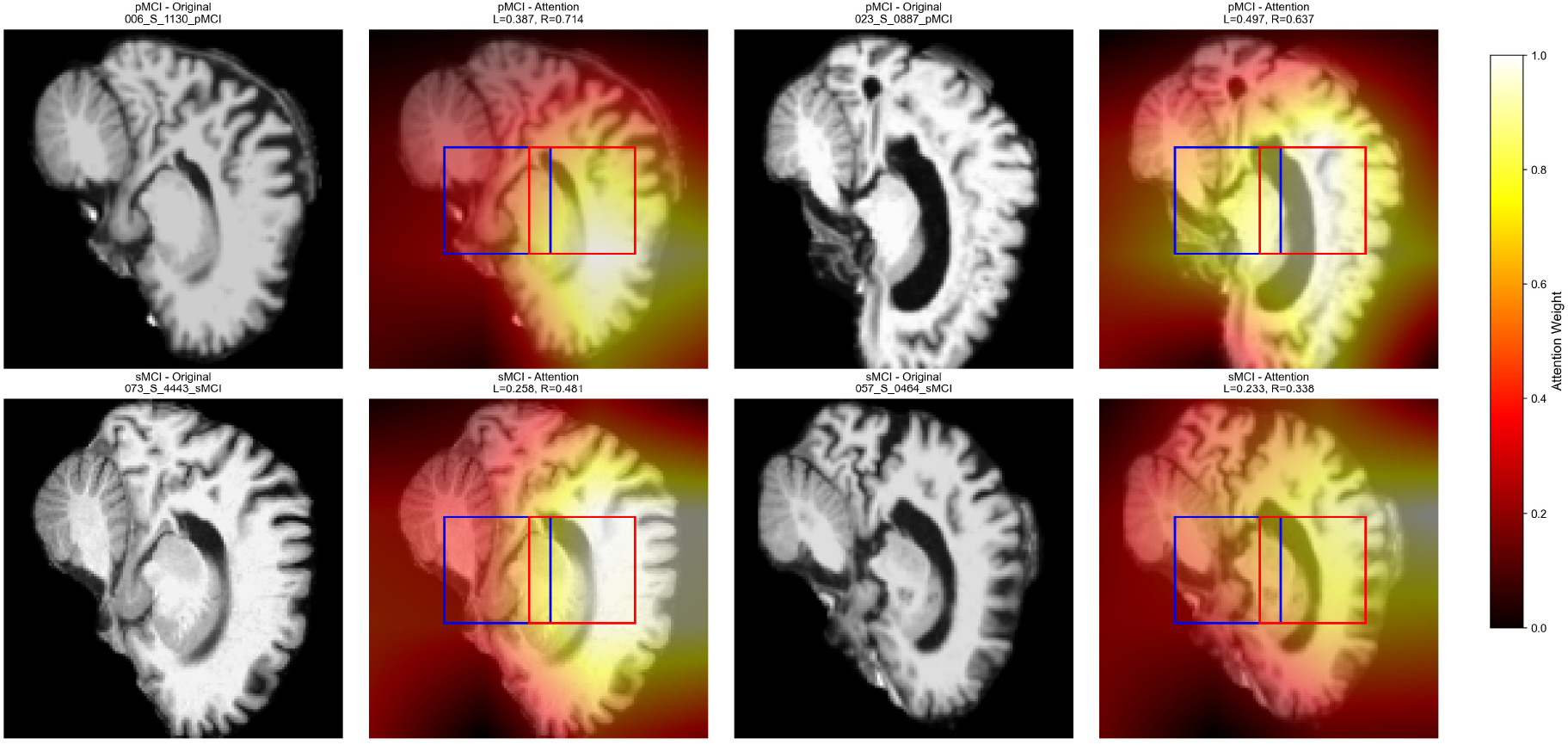
High-resolution spatial attention maps superimposed on anatomical MRI via trilinear upsampling. Red and blue boxes denote the right and left hippocampus, respectively. The heatmaps confirm a right-hemisphere dominance pattern, where pMCI subjects (top) exhibit significantly stronger activation in the right hippocampus (red box) compared to sMCI subjects (bottom), highlighting the model’s sensitivity to asymmetric pathology.

This asymmetry extends to the network level. SHAP value analysis (Fig. 12) identifies parietal asymmetry as the top predictor, led by the Angular Gyrus and SupraMarginal Gyrus. The distribution of positive SHAP values indicates that elevated right-dominant asymmetry drives predictions toward the pMCI class. The specific results are shown in Table 11. Differential analysis (Fig. 11) confirms that pMCI patients show exacerbated hemispheric asymmetry (Δ ≈ 0.011 in Angular Gyrus) compared to sMCI. This rightward shift in DMN hubs likely reflects the preferential vulnerability of left-hemisphere semantic nodes, resulting in compensatory or pathological lateralization toward the right hemisphere.

**Table 11.** Top-5 brain regions exhibiting hemispheric asymmetry differences.

| Rank | Region | Lobe | pMCI Asym. | sMCI Asym. | Diff. ($\Delta$) | Cohen's $d$ | p-value |
| --- | --- | --- | --- | --- | --- | --- | --- |
| 1 | Angular Gyrus | Parietal | 0.0865 | 0.0756 | 0.0109 | 0.174 | 0.046 |
| 2 | Supramarginal Gyrus | Parietal | 0.0853 | 0.0753 | 0.0100 | 0.135 | 0.120 |
| 3 | Middle Frontal Gyrus | Frontal | 0.0616 | 0.0535 | 0.0081 | 0.137 | 0.115 |
| 4 | Superior Frontal Gyrus | Frontal | 0.1106 | 0.1049 | 0.0057 | 0.117 | 0.180 |
| 5 | Orbitofrontal Cortex | Frontal | 0.0681 | 0.0633 | 0.0048 | 0.066 | 0.450 |

**Figure 10.**
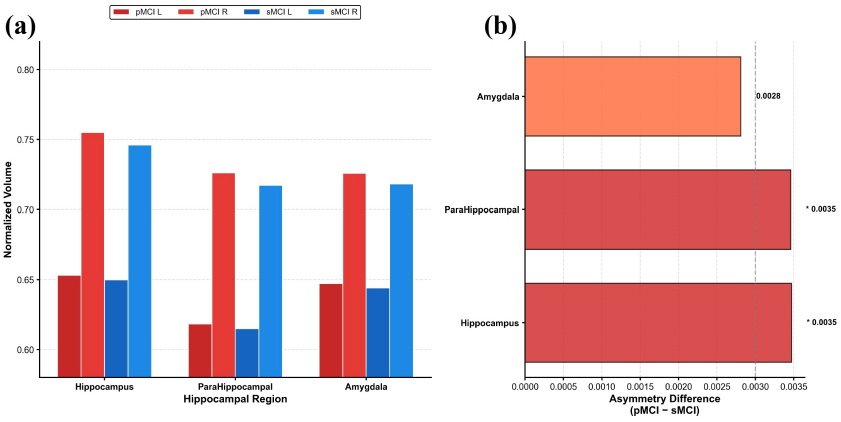
Hemispheric asymmetry of spatial attention in the medial temporal lobe. (a) Attention weights exhibit a consistent right-hemisphere dominance across hippocampal subregions. (b) Statistical analysis reveals significantly exacerbated rightward asymmetry in pMCI patients (*p* < 0.05), highlighting the vulnerability of spatial memory networks.

**Figure 11.**
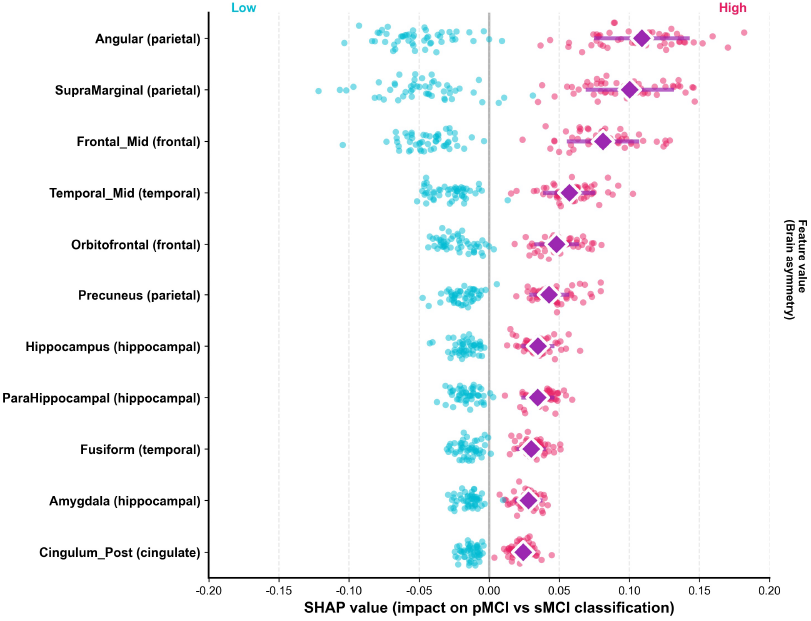
SHAP summary plot quantifying the contribution of hemispheric asymmetry to classification decisions. Parietal regions, particularly the Angular Gyrus, exhibit the highest impact. High feature values (red points), representing strong right-dominant asymmetry, correlate with positive SHAP values, thereby increasing the probability of pMCI.

**Figure 12.**
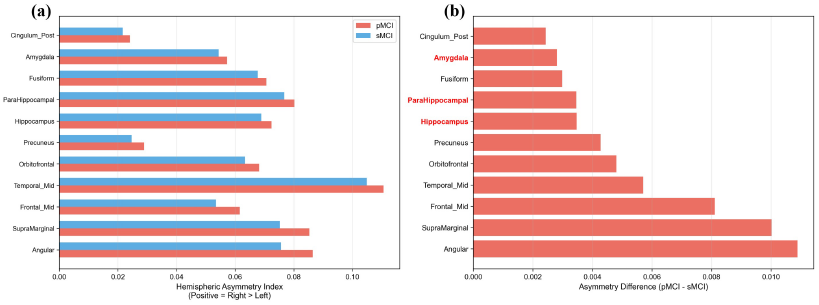
Quantitative analysis of hemispheric asymmetry. (a) pMCI subjects (red) exhibit elevated right-hemisphere dominance compared to sMCI across discriminative regions. (b) Group differences reveal exacerbated asymmetry in pMCI, with key medial temporal lobe structures highlighted in red.

Finally, unsupervised hierarchical clustering of attention patterns identified four Synergistic Impairment Patterns with high topological consistency (Fig. 13), validating the biological relevance of the model. Cluster 0 (High-Saliency DMN Hubs) captures core DMN nodes (Precuneus, Angular, PCC) with consistently high weights (> 0.8), serving as the primary discriminative basis. Cluster 2 (Right-Lateralized Compensatory Network) comprises right-hemisphere structures (e.g., R. Hippocampus) where elevated intensity confirms a critical compensatory role. In contrast, Cluster 1 (Vulnerable Left Limbic Subsystem) encompasses the Left Hippocampus and Amygdala, reflecting functional decline through reduced weights. Finally, Cluster 3 (Left Temporal Silent Zone) involves left parahippocampal regions with minimal weights, suggesting functional silencing or severe atrophy. This stepwise distribution, transitioning from left silencing to right compensation, validates the left decline-right compensation hypothesis at the network level, providing a granular basis for characterizing pMCI heterogeneity.

**Figure 13.**
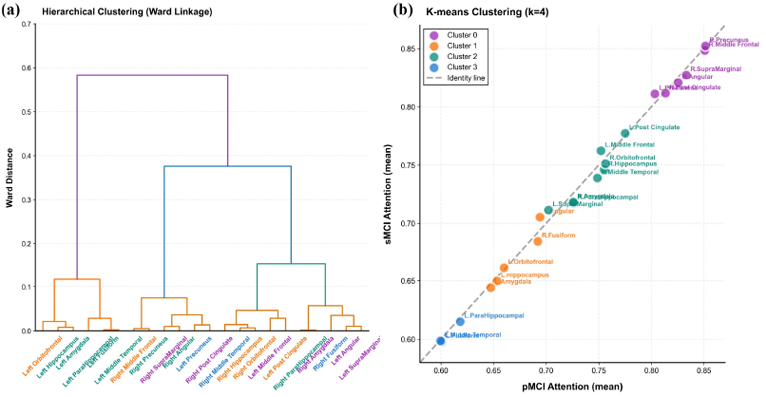
Identification of collaborative impairment patterns via unsupervised clustering. (a) Hierarchical clustering automatically groups 22 brain regions into anatomically distinct clusters. (b) K-means clustering (*k* = 4) reveals four distinct co-impairment patterns, providing data-driven insights into the clinical heterogeneity of pMCI.

## 5. Discussion

This study aims to provide a unified solution to the challenges of rigid modal fusion strategies and the passive utilization of clinical prior knowledge in early AD diagnosis. Beyond achieving performance breakthroughs in diagnostic tasks, our framework integrates conversion time prediction and survival analysis, establishing a comprehensive workflow for intelligent AD assessment. While multi-modal diagnosis has progressed, its capacity to capture subtle early pathological features remains behind clinical demands. Current methodologies often prioritize complex visual feature extractors or increasing modality count, while neglecting the active calibration of feature extraction using low-cost clinical priors. This oversight limits performance in high-difficulty tasks such as pMCI detection. By transforming clinical and cognitive states into spatial attention weights, the COMPASS framework instigates a paradigm shift from data-driven to clinical-prior-driven learning. This mechanism enables the model to emulate the awareness of a senior clinician, consciously focusing on specific brain regions based on cognitive scores, thereby mitigating noise from blind whole-brain searches and opening new perspectives for intelligent AD diagnostics.

On this basis, we propose the first clinical-guided multi-modal framework integrating diagnosis, conversion prediction, and survival analysis. Existing traditional methods for Late Fusion often assume image features are static, ignores the semantic gap between heterogeneous data, and hinders deep cross-modal mappings. Addressing this, we demonstrate through hierarchical ablation experiments that utilizing CGSA and DSAMF modules to position clinical data as antecedent guiding signals significantly improves sensitivity to subtle lesions, even without expensive advanced encoders. The core mechanism is that clinical guidance compels the network to attend to pathological regions such as the hippocampus and medial temporal lobe, effectively enhancing the signal-to-noise ratio in the feature space. This provides important design insights for future multi-modal medical image analysis models.

Concurrently, we quantified the neurobiological interpretability behind the model’s decisions. Unlike previous studies limited to qualitative heatmap visualization, we analyzed the distribution of attention weights across brain regions. Experiments demonstrate that the right hippocampus and medial temporal lobe exhibit a significant Right Hemisphere Dominance in early AD conversion prediction, challenging the traditional view of uniform bilateral atrophy. We suggest this occurs because the spatial memory network dominated by the right hippocampus is more vulnerable in the pMCI stage than the left-sided linguistic memory network, explaining why spatial disorientation is often an initial symptom. We also observed a Left Decline-Right Compensation pattern, indicating that when left hemisphere networks are impaired, the brain mobilizes right hemisphere resources to maintain cognitive homeostasis. These results confirm that the COMPASS model learns a physiologically meaningful pathological topology rather than memorizing data noise, effectively avoiding overfitting.

We must also emphasize the limitations of our method. First, the reliance on ADAS scores as a key guiding feature restricted validation primarily to the ADNI dataset, as this modality is frequently absent in other public cohorts. Second, the requirement for fully paired multi-modal data, specifically PET, necessitated the exclusion of incomplete samples, limiting the scale and diversity of training data. Future research will focus on leveraging unpaired data to extend the model’s generalization boundaries.

## 6. Conclusion

Pathological heterogeneity and the multi-modal semantic gap present significant barriers to early AD diagnosis, further complicated by the limited utilization of clinical priors in deep learning. Addressing these challenges, our COMPASS framework demonstrates that transforming clinical cognitive states into visual guiding signals is decisive for capturing subtle pathology. This approach not only achieves breakthroughs in diagnosis and time-to-conversion prediction but also harmonizes high-precision prediction with biological interpretability through the discovery of Right Hippocampal Dominance. Future work will integrate longitudinal imaging and broader modalities to advance the model’s utility in real-world clinical settings.

## Data Availability

Data used in this study were obtained from the Alzheimer s Disease Neuroimaging Initiative (ADNI) database. ADNI data are available to qualified researchers through the LONI Image and Data Archive (IDA) after application and approval, subject to the ADNI Data Use Agreement.

https://adni.loni.usc.edu/data-samples/adni-data/

## Data availability

The authors do not have permission to share data.

## Acknowledgments

This work was supported by grants from STI 2030—Major Projects (no. 2022ZD0209100); Natural Science Foundation of China (no. 82571771) and Natural Science Foundation of Shanghai (no: 25ZR1401167).

